# Cell-type-resolved genetic regulatory variation shapes inflammatory bowel disease risk

**DOI:** 10.1101/2025.06.24.25330216

**Authors:** Tobi Alegbe, Bradley T Harris, Laura Fachal, Lucia Ramirez-Navarro, Marcus Tutert, Monika Krzak, Mennatallah Ghouraba, Michelle Strickland, Matiss Ozols, Celeste E Cohen, Saniya Khullar, Eleonora Khabirova, Nikolaos I Panousis, David Ochoa, Noor Wana, May Xueqi Hu, Jason Skelton, Jasmin Ostermayer, Kimberly Ai Xian Cheam, D Leland Taylor, Yong Gu, Claire Dawson, Tina Thompson, Kenneth Arestang, Nilanga Nishad, Biljana Brezina, Charry Queen Caballes, Wendy Garri, Steven Leonard, Vivek Iyer, Miles Parkes, Chris Wallace, Rebecca E McIntyre, Cristina Cotobal Martin, Gareth-Rhys Jones, Tim Raine, Carl A Anderson

## Abstract

Most genetic variants associated with complex diseases lie in non-coding regions, complicating efforts to identify effector genes and relevant cell types. Here, we map cis-eQTLs across 2.2 million single cells from blood and intestinal biopsies of 421 individuals, including 125 with inflammatory bowel disease (IBD). Cell-type-level eQTLs were more distal to transcription start sites, enriched in enhancers, less likely to regulate the nearest gene, and over two-fold more likely to colocalise with IBD GWAS loci than eQTLs detected at tissue-level resolution. We nominate effector genes at over half of known IBD loci, including *MAML2*, *PSEN2,* and *ZMIZ1* in myeloid cells, implicating reduced Notch signalling in intestinal immune dysfunction. We also identify Wnt regulated genes, including *MYC*, in epithelial stem and progenitor cells, suggesting that impaired renewal contributes to barrier breakdown. Our results provide a mechanistic map linking genetic risk to specific genes and cell types in IBD, and a framework for effector gene discovery in complex disease.

## Introduction

Common complex diseases cause substantial morbidity and mortality worldwide, yet the biological mechanisms underlying susceptibility remain incompletely understood. Genetic variation contributes considerably to disease susceptibility, and genome-wide association studies (GWAS) have now identified hundreds of thousands of associations between genetic variants and complex diseases and traits^1^. However, over 90% of GWAS signals map to non-coding regions of the genome^2^, complicating efforts to identify dysregulated genes, pathways and cell types.

Non-coding variants are thought to influence disease risk by altering expression of nearby genes. This has prompted large-scale efforts to map cis expression quantitative trait loci (cis-eQTLs) and integrate them with GWAS findings. Although these studies have advanced our understanding of disease biology^3–7^, most GWAS loci remain unannotated. This lack of functional resolution poses a major challenge for therapeutic development, as drug targets supported by human genetic evidence are substantially more likely to progress successfully through clinical trials^8–11^. One possible explanation for this annotation gap is that eQTL and GWAS signals often have different genomic contexts: eQTLs are more frequently found in promoters, whereas GWAS signals are enriched in enhancers^12^. However, this divergence may reflect methodological biases in current eQTL discovery efforts, which often rely on low-resolution bulk RNA sequencing of heterogeneous tissues or coarsely sorted cell populations^13–16^. These approaches may favour detection of regulatory effects shared across multiple cell types, which are more likely to reside in promoters rather than enhancers^17^.

Single-cell RNA sequencing (scRNA-seq) can capture gene-expression variation at much higher cellular resolutions, enabling the identification of cell-type-specific eQTLs (sc-eQTLs) that may be inaccessible to bulk tissue studies^7,18–23^. However, it remains unclear to what extent eQTLs identified at different cellular resolutions improve effector gene nomination at GWAS loci.

We hypothesised that GWAS signals would be preferentially enriched among cell-type-specific sc-eQTLs. Variants that perturb gene expression broadly across multiple cell types may be more strongly constrained by negative selection, preventing them from reaching allele frequencies detectable by GWAS. In contrast, variants acting in a restricted set of cell types or contexts may face weaker selective pressures, allowing them to persist at higher frequencies and thus be captured by GWAS. Such variants would also be more likely to map to enhancers rather than promoters, aligning their genomic location more closely with GWAS signals.

To test this hypothesis, we established the IBDverse project, generating large-scale scRNA-seq datasets from blood and intestinal biopsies of the terminal ileum and rectum — the sites most commonly affected in Crohn’s disease (CD) and ulcerative colitis (UC), respectively. These diseases represent the two main forms of inflammatory bowel disease (IBD), a debilitating group of gastrointestinal disorders affecting over 4.9 million people worldwide. Using nearly 2.2 million single-cell transcriptomes from over 400 individuals, including 125 with IBD, we comprehensively mapped eQTL effects across a broad range of cellular resolutions, from individual cell types to tissue-level groupings. Our analyses enabled the nomination of candidate effector genes and cell types for over half of the 320 known IBD loci, including 74 loci where an effector gene is nominated for the first time. Furthermore, we systematically quantified the contribution of eQTLs detected at different cellular resolutions to GWAS association signals, demonstrating that cell-type-level eQTLs are substantially enriched among IBD GWAS loci and preferentially reside in distinct genomic contexts compared to eQTLs detected across broader cell groupings. These findings highlight the power of single-cell resolution for interpreting complex disease genetics and provide a general framework for effector gene discovery across diverse diseases.

## Results

### A single-cell atlas of IBD-relevant intestinal tissue and blood

To enable large-scale sc-eQTL mapping at sites relevant to inflammatory bowel disease (IBD), we created ‘IBDverse’ - single-cell RNA sequencing data generated from 275 healthy rectal biopsies, 119 Crohn’s disease (CD) terminal ileal biopsies, 243 healthy terminal ileal biopsies, and 95 blood samples from individuals with CD (Fig. 1a). CD patients were required to have ileal involvement, and the ileal biopsies included both inflamed and uninflamed tissue. After stringent quality control, we obtained over 1.8 million high-quality cells, which we clustered into 86 transcriptionally distinct groups across 9 major cell populations (Fig. 1b; Supplementary Fig. S1). Samples from all anatomical sites were analysed jointly to ensure consistent cell type assignment and to capture both site-specific and shared cell states. Following training and annotation with CellTypist^24^ (Methods), our final sc-eQTL mapping cohort comprised nearly 2.2 million cells from 397 individuals with matched genome-wide genotype data (Fig. 1c; Supplementary Fig. S2).

**Figure 1:**
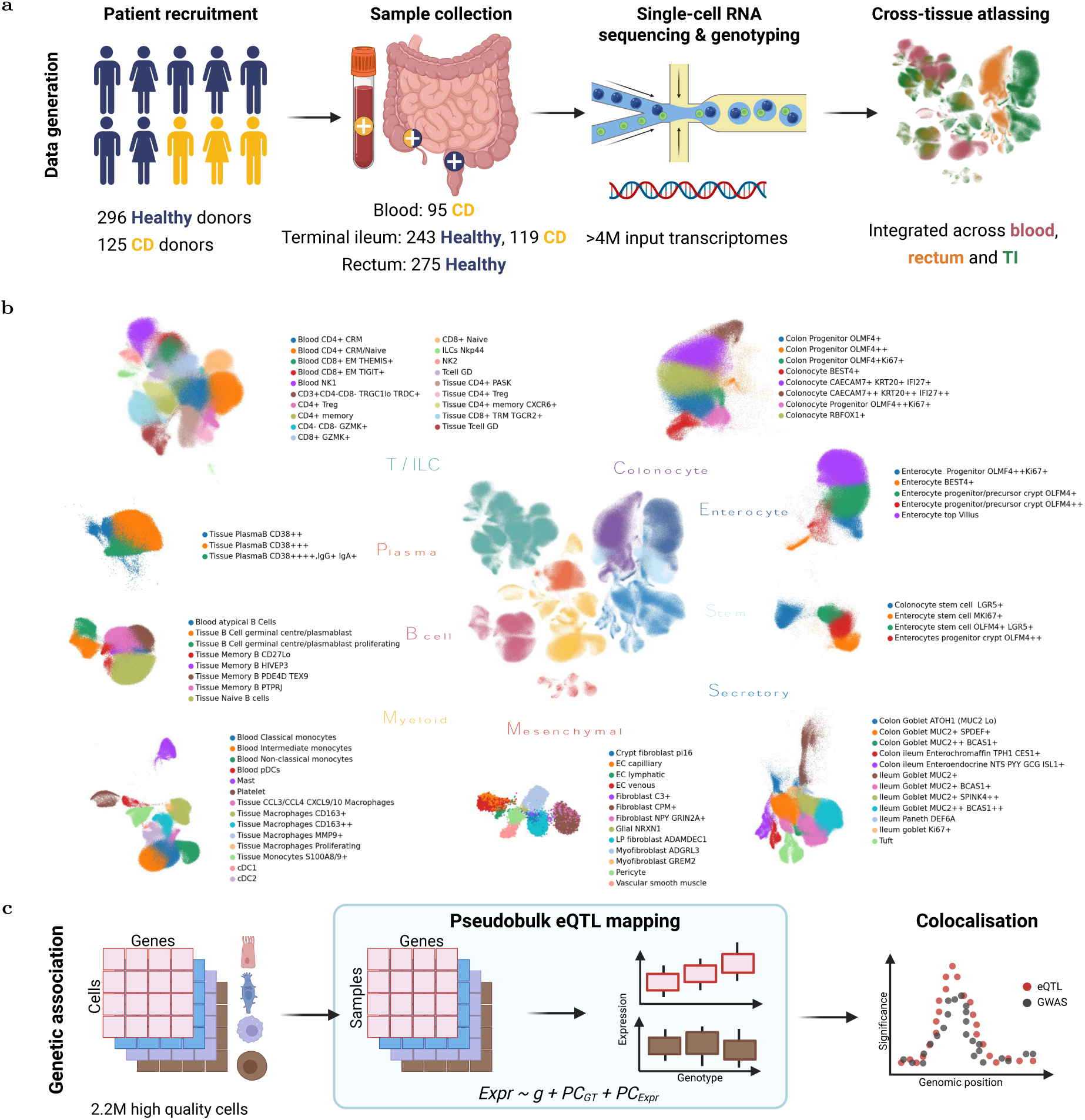
Mapping cells and sc-eQTLs across IBDverse. a) Single-cell RNAseq data was generated from blood, rectal, and terminal ileal (TI) biopsies obtained from 421 healthy individuals or CD patients. Quality control, integration and clustering was performed in a single analysis to generate a multi-site single-cell atlas of inflammatory bowel disease (see Methods, Supplementary Figure S1). Alongside this data, genome-wide germline genotype data was obtained from the same individuals. b) Uniform Manifold Approximation and Projection (UMAP) of the 9 major populations and 86 cell types identified after quality control and clustering. c) Pseudobulk cis-eQTL mapping was performed on 2,196,874 confidently annotated cells (see Methods). Sc-eQTLs were colocalised with disease susceptibility GWAS loci to nominate disease effector genes.

### Cell-type resolution reveals distinct eQTLs missed in bulk analysis

Leveraging the single-cell and multi-site nature of IBDverse, we performed cis-eQTL mapping across a broad range of cellular resolutions. Specifically, we mapped eQTLs at (1) the level of 86 transcriptionally defined cell types, (2) within 9 major cell populations, and (3) across all cells combined from each sample (’All cells’ resolution). This was done within each anatomical site separately (rectum, terminal ileum, or blood), or by combining expression across sites (‘cross-site’) (Fig. 2a), resulting in a theoretical maximum of 384 unique cellular annotations. Where eQTL effects were detected, we mapped conditionally independent signals and grouped lead variants based on linkage disequilibrium (LD) to enable direct comparison of eQTL effects across annotations and resolutions (Methods).

**Figure 2:**
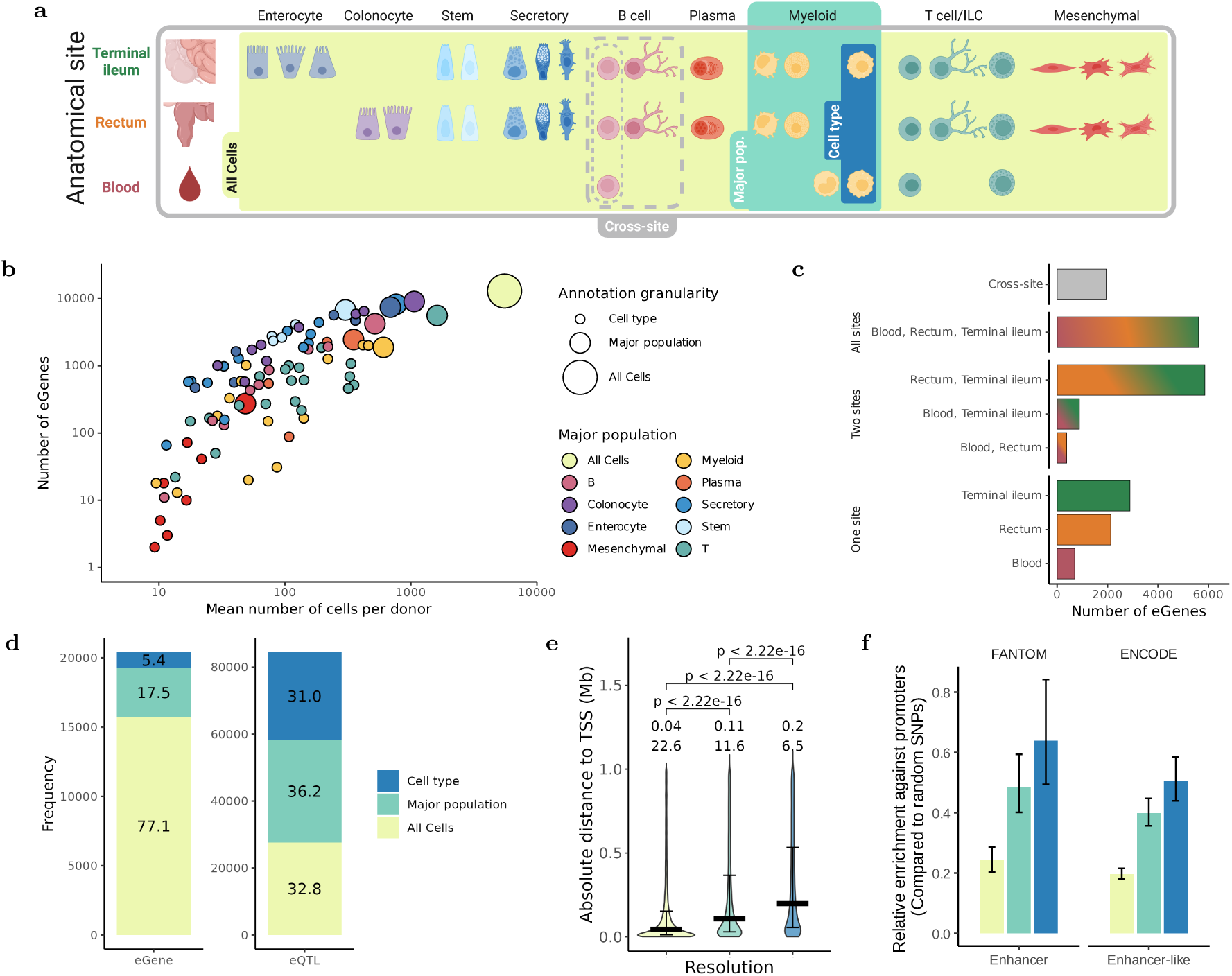
Distinct eQTLs are observed at different cellular resolutions. a) Schematic to illustrate the different axes by which cellular expression was aggregated for eQTL mapping; 1) by varying resolution - expression was aggregated at the level of individual cell-types (highest resolution), coarse major populations (middle resolution) or ignoring all annotations to aggregate all cells per sample, known as ‘All Cells’ (lowest resolution), and 2) by varying site - aggregating expression within each site individually or by combining sites together (’cross-site’). b) Association between the mean number of cells each donor contributed to each annotation and the number of eGenes detected in the cross-site analysis (logarithmised on both axes). c) The sharing of eGenes across sites, or identification in the cross-site analysis only. d) The identification of eGenes (left) and eQTLs (right) at different resolutions. Genes and eQTLs are grouped into the minimum resolution at which they were detected. The percentage of effects found at each resolution is displayed. e) Distance from the lead variant of each eQTL to the transcription start site (TSS) of their associated eGene, grouped by the minimum resolution at which a given eQTL was detected as in (d). The median distances per group (megabases, Mb) and percentage of eQTLs located less than 10kb from the TSS of their associated eGene are also displayed. Colour legend is shared with (d). f) Relative enrichment of eQTLs detected at each resolution in FANTOM and ENCODE enhancer genomic annotations over promoters. Error bars represent the 2.5 and 97.5 percentile from bootrapping sampled variants (see Methods). P-values for two-sided t-tests within annotations were all < 2.2e-16. Colour legend is shared with (d).

We identified 84,376 eQTLs regulating the expression of 20,389 of the 24,761 genes tested (82.2%). 252 of the possible 384 cellular annotations had a sufficient number of samples for testing and at least one eQTL was detected in 251 out of these 252 annotations (Methods), with the number of eGenes (genes with at least one significant eQTL) being positively correlated with the number of constituent cells, the number of donors, and the average number of genes expressed per cell (all p < 0.001; Fig. 2b; Supplementary Fig. S3). Most eGenes were shared across sites, most frequently between the rectum and terminal ileum (5,861; 28.7%), followed by all three sites (5,613; 27.5%) (Fig. 2c). However, many eGenes (1,945, 9.5%) were only detected when combining sites, displaying the value of aggregating common cell types across different tissues.

The majority of eGenes (15,713; 77.1%) were detected at the ‘All cells’ resolution, with additional eGenes identified at the major population (3,565; 17.5%) and cell-type levels (1,111; 5.4%) (Fig 2d). In contrast, nearly one third (26,192; 31%) of eQTLs were detected at only the cell-type level, with even more detected at the major population but not ‘All cells’ level (30,540; 36.2%). The number of eQTLs per eGene also declined at coarser resolutions—averaging 3.1 at the cell-type level, 2.7 at major population level, and 1.9 at the ‘All cells’ level (all comparisons p < 2.2 × 10⁻¹⁶; Supplementary Fig. S4). Together, these findings demonstrate a key distinction: while the ‘All cells’ resolution identifies more eGenes overall, cell-type-level mapping reveals distinct and diverse regulatory variants. High-resolution eQTL mapping therefore uncovers regulatory effects that are masked in bulk analyses.

Given these differences in detection, we next asked whether the genomic location of eQTLs also varied by resolution. eQTLs detected only at the cell-type level were located significantly further from the transcription start site (TSS) of their associated gene (median distance = 200 kb) than those detected at the major population (median = 108 kb) or ‘All cells’ level (median = 44 kb; both comparisons p < 2.2 × 10⁻¹⁶; Fig. 2e). Correspondingly, the nearest gene was the eGene for 38.3% of eQTLs detected at the ‘All cells’ level, compared to 28% and 20.1% for those at the major population and cell-type levels, respectively (Supplementary Fig. S5).

Functionally, eQTLs identified at only the cell-type level were more enriched in FANTOM and ENCODE enhancer annotations rather than promoters, compared with those at the ‘All Cells’ or major population level (p<2.2e-16 for all comparisons) (Fig. 2f). Because GWAS loci are also enriched in enhancers rather than promoters^12^, these findings suggest that eQTLs detected at higher transcriptional resolution may more closely align with the regulatory architecture of complex trait associations.

We next asked whether genetic effects on gene expression are modulated by individual-level traits. We performed interaction eQTL (ieQTL) mapping using genotype–expression models that included interaction terms for age, sex and smoking status as well as Crohn’s disease diagnosis and degree of ileal inflammation, where possible (Methods). We identified 1,051 ieQTLs regulating the expression of 977 genes (ieGenes), and 764 (78.2%) of these ieGenes were modulated by gut inflammation status. Strikingly, the majority of these inflammation-responsive ieGenes (501; 65.6%) were detected in enterocytes, suggesting that genetic effects on gene expression in this major population are highly sensitive to inflammatory context (Supplementary Fig. S6).

Although most ieGenes (941; 96.3%) were also eGenes, only 44 ieQTLs (4.2%) were in strong LD with eQTLs (r^2^>0.5), indicating these effects are largely distinct from those observed in conventional eQTL analyses. To better understand the pathways disrupted by the interaction between inflammation and genetics in enterocytes we performed gene set enrichment analysis, which returned 4 pathways (coefficient > 0; FDR < 0.05; Supplementary Material S1). Notably, the most enriched pathway was ‘response to metal ion binding’ in enterocytes (coefficient = 2.04); expression of the leading edge genes have previously been shown to be protective against intestinal inflammation in mice^25,26^. These findings demonstrate that genetic variation contributes to inter-individual differences in cellular responses to inflammation, and that interaction eQTL mapping using scRNA-seq data can identify context-dependent regulatory mechanisms relevant to disease.

### Cell-type-level eQTLs more effectively nominate disease effector genes

Having generated a comprehensive atlas of cis-regulatory effects across disease-relevant tissues, cell types, and inflammatory contexts, we hypothesised that this resource would provide a powerful substrate for identifying effector genes at IBD GWAS loci. In particular, we reasoned that cell-type-specific eQTLs—being more distal to transcription start sites and enriched in enhancer elements—would be more likely to colocalise with GWAS signals than eQTLs identified at coarser resolutions.

We therefore performed colocalisation analyses between IBD GWAS signals and our full set of eQTL and interaction eQTL (ieQTL) maps, using summary statistics for Crohn’s disease (CD), ulcerative colitis (UC), and a combined IBD phenotype^3^. To benchmark our findings, we compared them to colocalisations previously reported for IBD loci in the Open Targets Genetics portal, including those based on expression, splicing, and other molecular QTLs^27^.

Among the 321 IBD loci reported to date^28^, we observed colocalisation (posterior probability of a shared causal variant, PPₕ₄ > 0.75) with either eQTLs or ieQTLs at 180 loci (56%), implicating 419 distinct effector genes (Table 1; Supplementary Material S2). This includes 74 loci where no effector gene had previously been nominated by colocalisation in the Open Targets Genetics portal (Fig. 3a). Notably, at 2 of these 180 loci, colocalisation was observed only with ieQTLs, illustrating that context-dependent regulatory effects can reveal additional candidate effector genes not detected under baseline conditions.

**Figure 3:**
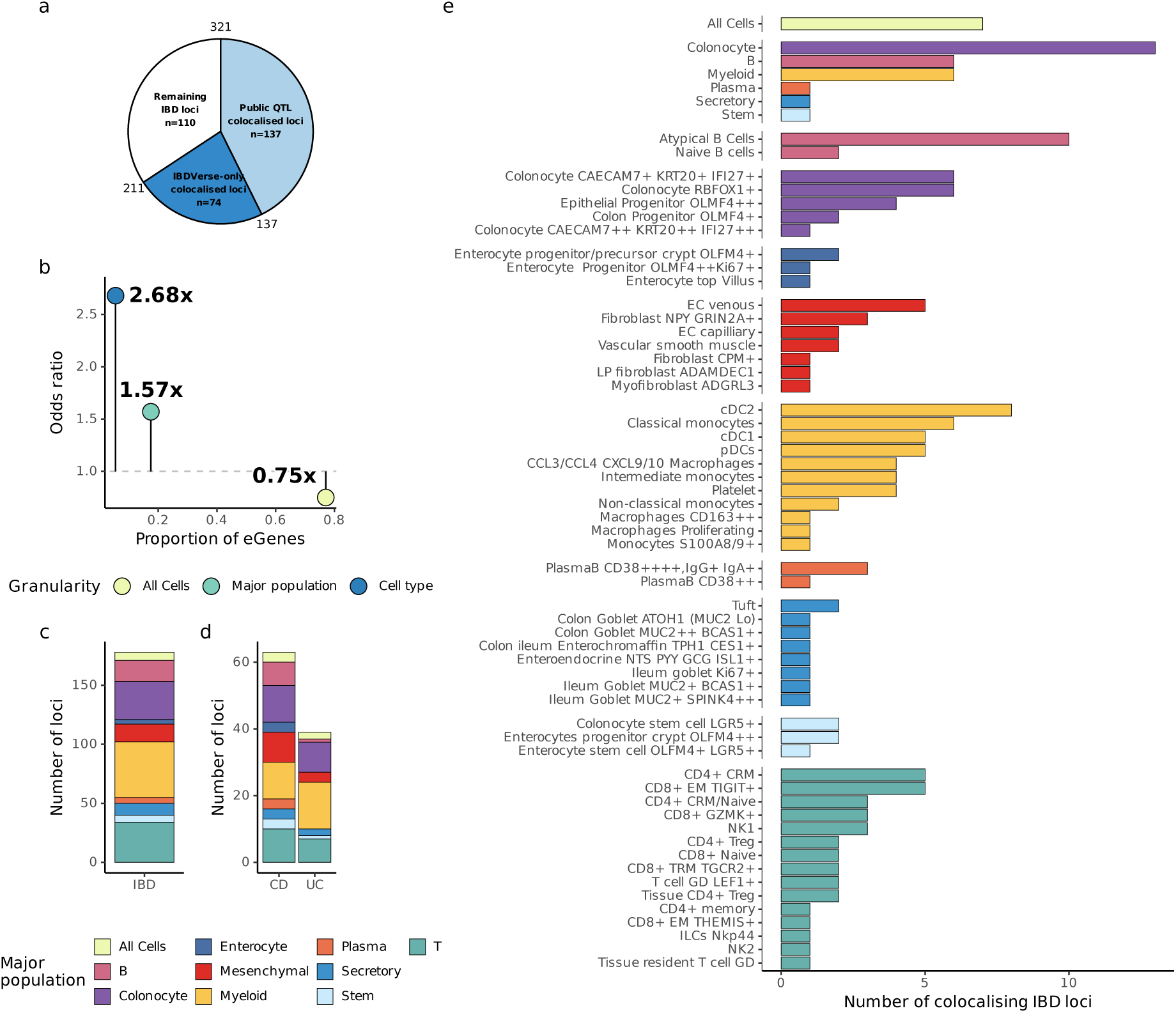
Detection and distribution of colocalisation events with IBD, CD and UC GWAS. a) The number of IBD GWAS loci (Liu et al. 2023) coloured by the proportion that previously colocalised (posterior probability of hypothesis 4; PPH4 > 0.75) with existing bulk QTL studies in Open Targets Genetics (see Methods), or do so for the first time with IBDverse sc-eQTLs. b) The proportion of eGenes, grouped by the lowest resolution they can be detected (x-axis) against the odds-ratio between eGene detection disease effector gene detection at each resolution (y-axis). c) The number of IBD, or subtype-specific (d), GWAS loci that colocalise with IBDverse eQTLs (PPH4 > 0.75), coloured by the major population (or All Cells) where the variance in disease effector gene expression explained by the colocalising variant was greatest. (e) The number of loci where variance in disease effector gene expression explained by the colocalising variant was greatest across cell-types. Bars for each annotation are coloured by the major population they belong to, following the key of (c,d).

**Table 1:** Novel disease effector genes of high confidence. IBD loci for which we identify a high confidence colocalisation (PPH4 > 0.9) between dysregulation of a single gene with IBD, CD or UC susceptibility, and no disease effector gene has been nominated at this locus by colocalisation previously in the Open Targets Genetics database. Extended summary statistics for all loci are available in Supplementary Material S2. GWAS lead = lead variant for GWAS signal after intersection with variants tested for eQTL. Maximum PPH4 = maximum posterior probability of colocalisation observed for given eQTL and the GWAS signal in any condition. The primary GWAS signal in locus 300 is driven by rs8176719 (chr9:133257521:T:TC - p.Thr87AspfsTer107 – ABO type 0*) so we cannot fully disregard that the colocalisation we observe is mediated by the coding variant.

|  | Locus ID | GWAS trait | GWAS lead | Gene symbol | Interaction | Maximum PPH4 |
| --- | --- | --- | --- | --- | --- | --- |
| 1 | 9 | CD, IBD | chr1_77984833_C_A, chr1_78381145_G_A | FUBP1 | base | 1.00 |
| 2 | 56 | UC | chr3_46410189_A_G | CCR1 | base | 0.92 |
| 3 | 70 | UC | chr4_73998280_T_C | MTHFD2L | ses_cd_binned | 0.92 |
| 4 | 72 | UC, IBD | chr4_105173270_G_A, chr4_105160479_T_C | TET2 | base | 0.94 |
| 5 | 91 | IBD | chr5_172900358_T_C | STK10 | base | 0.91 |
| 6 | 101 | IBD, CD, UC | chr6_42039665_C_T, chr6_42040465_C_G | CCND3 | base | 1.00 |
| 7 | 124 | IBD, UC | chr7_117254872_A_G, chr7_117252792_T_C | CTTNBP2 | base | 0.91 |
| 8 | 133 | CD, IBD | chr8_128540294_G_A | MYC | base | 0.92 |
| 9 | 166 | UC, IBD | chr11_96298708_T_A | MAML2 | base | 0.98 |
| 10 | 172 | IBD, CD | chr12_40220632_C_T, chr12_40430937_G_A | LRRK2-DT | base | 0.92 |
| 11 | 173 | UC | chr12_47814585_T_C | VDR | base | 0.91 |
| 12 | 175 | IBD, CD, UC | chr12_111427245_C_G, chr12_111446804_T_C | ARPC3 | base | 0.94 |
| 13 | 176 | CD, IBD | chr12_119709120_G_A | PRKAB1 | base | 0.97 |
| 14 | 178 | UC | chr13_40252229_G_A | FOXO1 | base | 0.97 |
| 15 | 184 | IBD, CD | chr13_99348586_A_T, chr13_99337284_A_G | GPR183 | base | 0.91 |
| 16 | 188 | CD | chr15_38608111_C_A | RASGRP1 | base | 0.97 |
| 17 | 214 | IBD, CD | chr19_1123379_A_C, chr19_1123653_G_A | REXO1 | base | 0.97 |
| 18 | 225 | CD | chr20_44210072_C_G | OSER1 | base | 0.91 |
| 19 | 233 | CD, IBD | chr21_44001056_A_G | GATD3 | base | 0.95 |
| 20 | 238 | IBD, CD, UC | chr22_39359768_C_T, chr22_39352907_G_A | ADSL | base | 0.95 |
| 21 | 271 | UC | chr1_205770138_C_T | CDK18 | base | 0.98 |
| 22 | 292 | UC | chr7_46834111_A_T | IGFBP3 | base | 0.97 |
| 23 | 300 | IBD, CD | chr9_133256625_T_C | ABO | base | 0.97 |
| 24 | 312 | IBD, CD, UC | chr12_69356604_A_T, chr12_69382403_A_G | YEATS4 | base | 0.94 |
| 25 | 335 | IBD | chr18_50918898_G_A | ME2 | base | 0.90 |
| 26 | 340 | CD | chr20_14563980_G_A | FLRT3 | base | 0.97 |

Our eQTLs and ieQTLs also colocalised with 104 of the 138 (75.6%) GWAS loci already reported to colocalise with molecular QTLs in the Open Target Genetics portal. At 78 of these loci, at least one effector gene colocalised in both our data and the Open Targets analysis. Notably, 103 of the previously implicated genes had only been supported by colocalisation in non-gut tissues, whereas our data revealed colocalisation in gut tissue for the first time. For 29 of the overlapping loci (37.2%), colocalisation in our study was restricted to a single major cell population or anatomical site, allowing refinement of prior hypotheses to suggest the specific tissue or cell type in which the candidate effector gene may be mediating its effect on disease susceptibility. Together, these findings significantly extend effector gene nomination at IBD loci and enable more targeted hypotheses about the cellular contexts in which genetic risk of IBD is exerted.

We next asked whether eGenes identified at the cell-type, major population or tissue-level differed in their likelihood of colocalising with IBD GWAS loci. GWAS signals were significantly more likely to colocalise with eQTLs identified at the cell-type level (ratio of odds [OR] between eGene detection and disease-effector gene nomination = 2.68), with more modest enrichment for major population-level eQTLs (OR = 1.57) and negative enrichment for those identified at the ‘All cells’ level (OR = 0.75) (Fig. 3b). This pattern was not explained by cell-type-specific expression of genes at these loci, as expression specificity did not differ between eGenes that colocalised at different resolutions (minimum Wilcoxon p = 0.38; Supplementary Fig. S7). These findings support the hypothesis that disease-relevant regulatory variation is often active in restricted cellular contexts, and that mapping eQTLs at cell-type resolution increases power to detect the effector genes underlying GWAS associations.

To refine effector cell-type hypotheses, we identified, for each colocalised signal, the cell type in which the lead variant explained the greatest proportion of variance in gene expression (Methods). This approach mitigates power-related biases across eQTL maps that would affect interpretation if based solely on the cell type with the highest posterior probability of colocalisation. By focusing on variance explained rather than detectability, we aimed to better prioritise cellular contexts likely to mediate disease risk. Across all IBD loci with colocalising eQTLs, myeloid cell populations were most frequently the cell type in which the lead variant explained the largest proportion of gene expression variance (47 loci; 26.4%), followed by T/ILC cells (34 loci, 19.1%) and colonocytes (32 loci; 18%) (Fig. 3c; Supplementary Material S3). Broadly, enrichment in these three major populations held when stratifying by disease subtype; at CD-specific loci, the most likely effector cell types were myeloid cells and colonocytes (11 loci each; 17.5%) and T/ILC cells (10 loci; 15.9%), while in UC, they were most often myeloid cells (14 loci; 36%) and colonocytes (9 loci; 23.1%) (Fig. 3d).

### Dysregulation of Notch signalling in dendritic cells implicated in IBD susceptibility

Amongst myeloid cell types, dendritic cells (cDC2, and cDC1, and pDC), were three of the four cell types most frequently nominated as the likely effector cell types, with a total of 18 IBD loci showing the highest proportion of gene expression variance explained in these populations. In humans, conventional dendritic cells (cDCs) originate from CD34⁺ bone marrow progenitors and differentiate into transcriptionally and functionally distinct subsets, including cDC1 and cDC2. These subsets are classically defined by abundance of CD141 and CD1c proteins, and more recently by *XCR1* and *CLEC10A* expression, reflecting their reliance on IRF8 and IRF4, respectively^29,30^. While both contribute to antigen presentation, they differ in pathogen sensing, cytokine production, and T cell priming, particularly in mucosal tissues. These functions are unique to cDCs and essential for oral tolerance and T cell priming, placing them at a critical interface between innate and adaptive immunity in the gut. Their recurrent nomination as likely effector cell types at IBD loci suggests a central role for dendritic cell–intrinsic pathways in shaping disease risk.

Of the 13 IBD loci where the strongest colocalising regulatory effects were observed in cDCs, disease effector genes at two are associated with Notch signalling, including *MAML2*. This gene is widely expressed across major populations (Fig. 4a), and encodes a nuclear co-activator essential for canonical Notch pathway activation: following ligand-induced release of the Notch intracellular domain (NICD), MAML2 binds RBP-J and enables transcription of target genes such as *HES1*^31,32^. In our data, a *MAML2* eQTL colocalised with an UC GWAS signal in cDC2s from the TI (PPₕ₄ = 0.98), and was the only gene supported at that locus (Fig. 4b). This represents a novel colocalisation not reported in the Open Targets Genetics Portal. Although we identified eight distinct eQTLs for *MAML2* across cell types and tissues (Supplementary Fig. 8), colocalisation with UC was observed for a single eQTL effect that was in low linkage disequilibrium with the others (maximum r² = 0.3), suggesting the disease association arises from a distinct regulatory effect active in dendritic cells.

**Figure 4:**
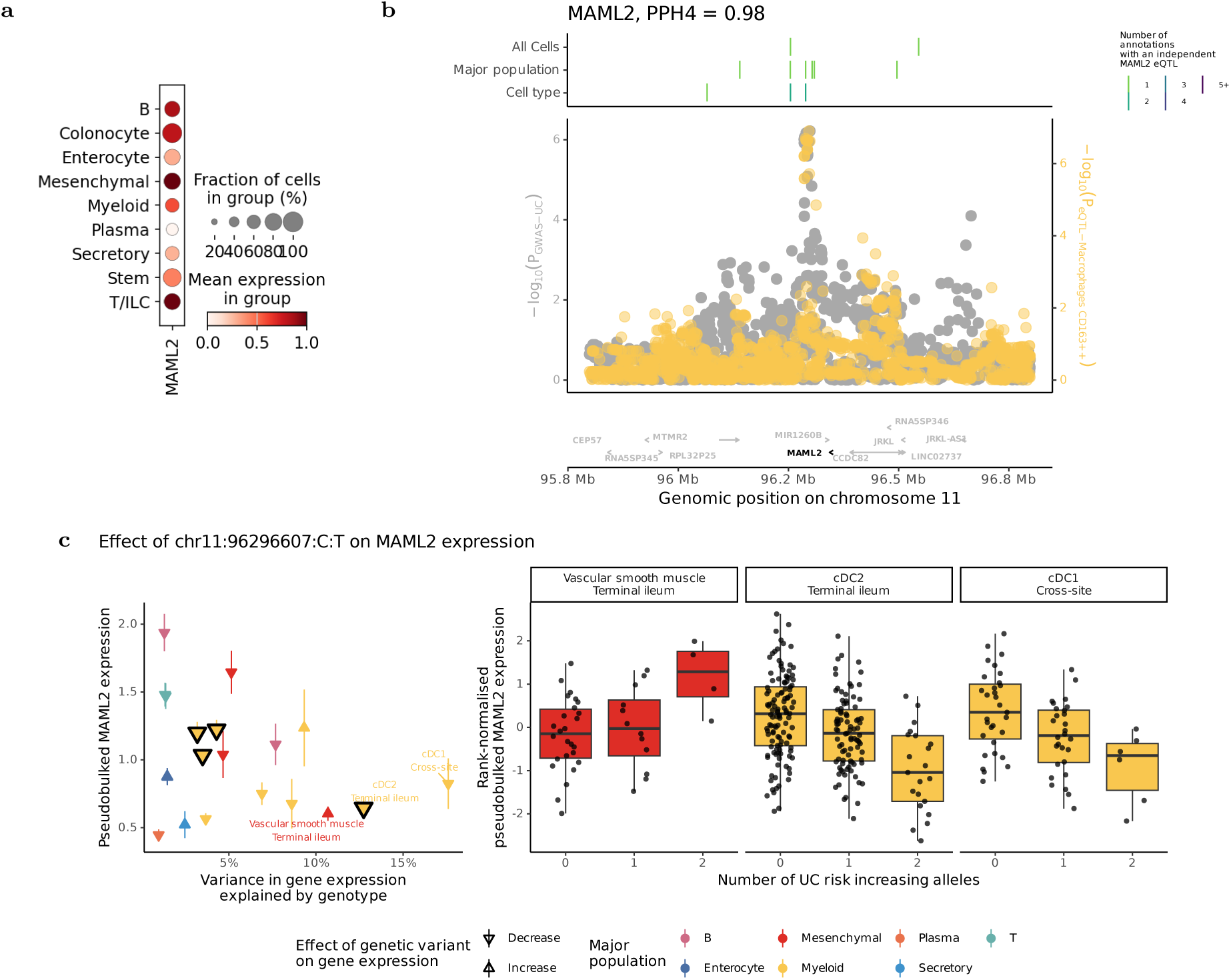
Novel colocalisation between *MAML2* dysregulation and IBD highlights susceptibility-associated dysregulaton of Notch signalling in conventional dendritic cells. (a) Expression of *MAML2* (log[1+counts per ten thousand]) across all major populations. (b) Significance of local variants for a genetic association with UC susceptibility (grey) and dysregulation of *MAML2* expression (yellow, following key of Fig. 3c). Boxes above indicate the position, resolution and number of annotations *MAML2* eQTLs can be identified. (c) Left - Median expression of *MAML2* and the amount of variance in normalised *MAML2* expression explained by genotype at the colocalising variant for each annotation with a nominal eQTL effect (p < 0.05) and median expression greater than zero. Annotations where colocalisation is detected (PPH4 > 0.75) are outlined. Right - The inverse normal transformed expression levels of *MAML2* in the three annotations where the greatest variance in gene expression is explained by this variant.

Although colocalisation was detected in cDC2s, the lead variant also explained substantial variance in *MAML2* expression in cDC1s (Fig. 4c). We interpret this as likely reflecting reduced power to detect colocalisation in cDC1s, which are both rarer in intestinal tissue and less well represented in our dataset. One possible concern is that colocalisation in dendritic cells may simply reflect high baseline expression of *MAML2* in these populations. However, we found no correlation between the strength of eQTL effects and baseline *MAML2* expression across cell types (Spearman’s R = –0.29; p = 0.21), suggesting that the disease association is driven by a cell-type-specific regulatory effect, not by general gene expression levels. Together, these results support the conclusion that dysregulation of *MAML2* expression by this specific sc-eQTL in cDCs contributes to UC risk.

An eQTL for an additional Notch pathway gene, *ZMIZ1,* also colocalised with IBD GWAS signals in dendritic cells (Supplementary Fig. 9). *ZMIZ1* encodes a transcriptional co-regulator of STAT and Notch pathways, and its deletion in mice phenocopies disruption of Notch ligand activity^33^. In our data, a variant associated with reduced *ZMIZ1* expression colocalised with CD risk (PPₕ₄ = 0.99), with the strongest effect observed in cDC2s from the rectum (23% variance explained). While we refer to these cells as cDC2s based on their transcriptional profile, we note that monocyte-derived DCs (mo-DCs) are indistinguishable from conventional DC2s in our atlas. Together with findings at *MAML2*, this result suggests that disruption of Notch pathway activity across dendritic cell subsets may play a protective role in maintaining intestinal homeostasis potentially by promoting tolerance or limiting pro-inflammatory signalling at the mucosal surface.

### Genetic variation implicates epithelial differentiation and Wnt signalling in IBD

We next turned to colocalisations implicating epithelial cells, where barrier dysfunction represents an established mechanism of IBD susceptibility. Colonocyte populations were among the most frequently nominated effector cell types, particularly the *RBFOX1⁺* and *CEACAM7⁺ KRT20⁺ IFI27⁺* expressing subsets (6 loci each), along with the broader colonocyte compartment at the major population level (13 loci) (Fig. 3e). At one such locus, we replicate a previously reported colocalisation between IBD risk and an eQTL associated with reduced expression of *FERMT1* (PPₕ₄ = 0.96, Supplementary Figure S10)^34–36^. Null variants in *FERMT1* are the most common defect in Kindler syndrome, a rare condition of epithelial fragility caused by lack of kindlin-1, which anchors the actin cytoskeleton to the plasma membrane, and is an important cause of monogenic IBD^37–39^. Our single cell analysis refines this colocalisation to epithelial populations, and *CEACAM7⁺ KRT20⁺ IFI27⁺* colonocytes in particular (Supplementary Material S3).

Several novel colocalisation events also implicated genes involved in epithelial differentiation and renewal. An eQTL for *RASGRP1* colocalised with CD risk (PPₕ₄ = 0.97), with the risk allele associated with increased *RASGRP1* expression in colonocytes (Fig. 5a, Supplementary Figure S10). *RASGRP1* encodes a Ras guanine nucleotide exchange factor that modulates MAPK signalling and epithelial proliferation^40,41^, and its depletion results in colitis phenotypes in mice^42^, suggesting a role in maintaining epithelial turnover. In addition, an eQTL for *LPIN3* colocalised with IBD risk (PPₕ₄ = 0.89), with the strongest effect observed in *CEACAM7⁺ KRT20⁺ IFI27⁺* colonocytes (Supplementary Material S3). *LPIN3* encodes Lipin 3, a phosphatidic acid phosphatase involved in lipid metabolism that also regulates Wnt signalling^43^, a pathway essential for maintaining epithelial stem cell renewal and mucosal integrity. Disruption of Wnt signalling can impair epithelial regeneration, promote barrier permeability, and drive intestinal inflammation^44,45^.

**Figure 5:**
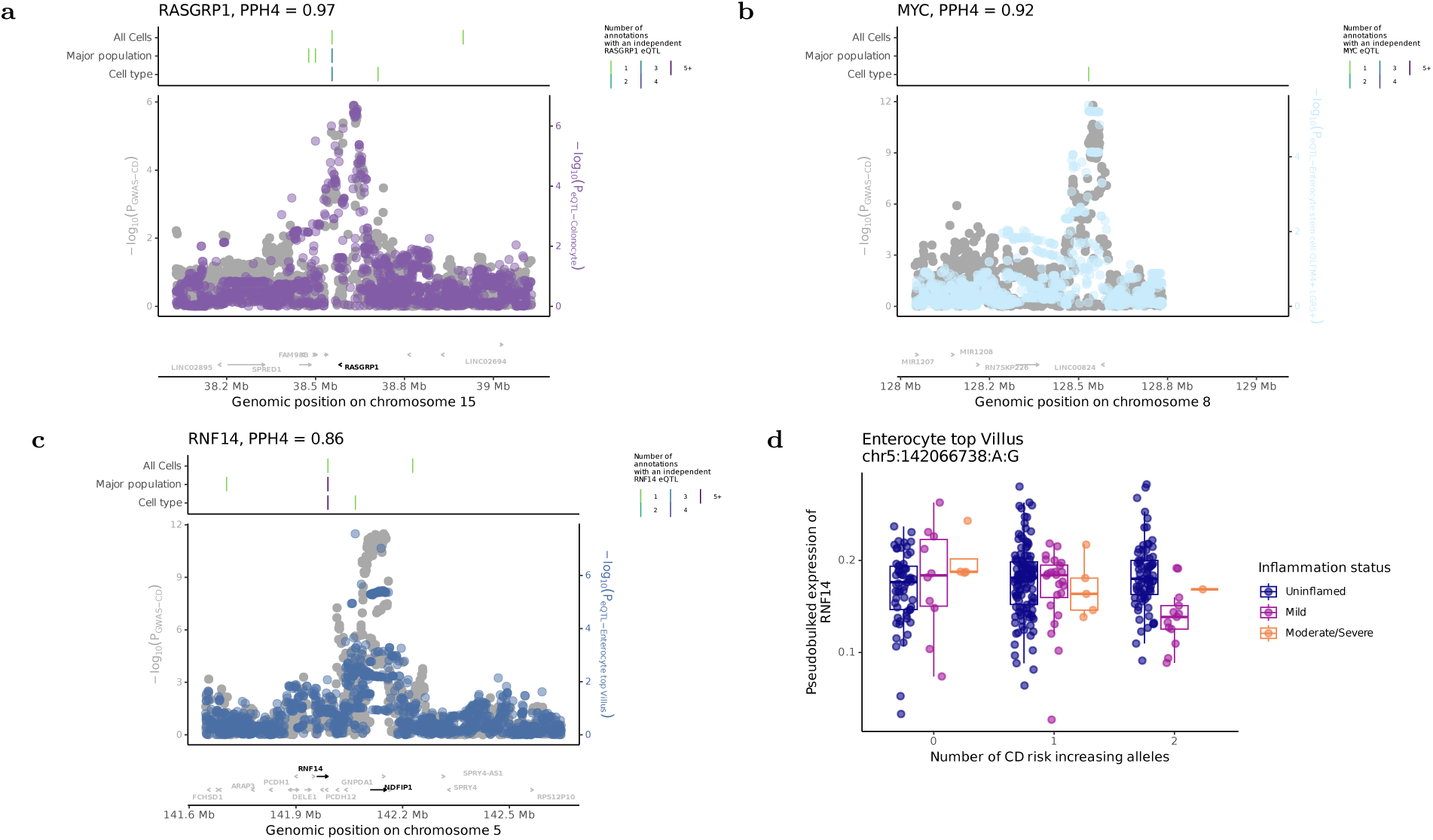
Wnt associated genes *RASGRP1*, *MYC* and *RNF14* colocalise with IBD susceptibility loci. (a-c) Local pattern of genetic association of variants for IBD GWAS susceptibility (grey) and dysregulation of *RASGRP1*, *MYC* and *RNF14* expression (non-grey, following previous colour keys of major populations). Lines above indicate the position, resolution and number of annotations distinct eQTLs for each gene can be identified in. (d) The interaction between genotype at the colocalising variant for *RNF14* expression in enterocytes from the top of the villu and disease severity.

Further supporting this mechanism, an eQTL regulating increased *MYC* expression colocalised with CD risk in *OLFM4⁺ LGR5⁺* intestinal stem cells (PPₕ₄ = 0.92, Fig. 5b, Supplementary Figure S10). *MYC* is a canonical Wnt target and master regulator of epithelial proliferation and metabolism, and its dysregulation perturbs intestinal homeostasis^46–48^. Importantly, *MYC* encodes c-Myc, a well-established oncogenic transcription factor that drives tumourigenesis through both somatic amplification and germline regulatory variation^49–52^, highlighting its dosage sensitivity and relevance to epithelial pathophysiology. A role for *MYC* expression in gastrointestinal inflammation is further supported by functional evidence from a DSS-induced colitis model, in which pharmacological activation of Wnt signalling improved disease severity and restored *Myc* expression in epithelial cells^53^.

We also identified colocalisation between CD risk and an eQTL for *FUBP1* (PPₕ₄=1.00, Supplementary Figure S10), a regulator of *MYC* and Wnt components such as *DVL1*^54,55^. Interestingly, the direction of effect varied by cell type: the IBD risk allele was associated with decreased *FUBP1* expression in *ADGRL3*-expressing myofibroblasts, where FUBP1 may support epithelial regeneration and mucosal repair^56^, but increased expression in several immune cell types, including T cells and rectal cDC2s (Supplementary Figure S10). These divergent regulatory effects suggest that *FUBP1* may influence IBD susceptibility through distinct mechanisms across cellular compartments. In immune cells, elevated *FUBP1* could plausibly enhance proliferative or metabolic programmes via *MYC* activation^57^ or modulate Wnt signalling dynamics^58^, while reduced expression in stromal cells may impair transcriptional programmes that support epithelial regeneration, including those involved in Wnt ligand production and mucosal repair.

An inflammation-dependent ieQTL regulating *RNF14* colocalised with CD susceptibility (PPₕ₄=0.86, Fig. 5c). *RNF14* encodes ring finger 14 protein, an E3 ubiquitin ligase that enhances Wnt signalling by stabilising TCF/LEF transcription factors, thereby promoting the expression of downstream targets, including *MYC*^59,60^. In inflamed enterocytes, the CD risk allele was associated with reduced *RNF14* expression (Fig. 5d), suggesting that Wnt signalling and epithelial regeneration may be impaired in the presence of inflammation. However, because our power to detect inflammation-dependent eQTLs outside of enterocytes is limited, we cannot exclude the possibility of larger effects in other cell types.

Together, these findings nominate dysregulation of epithelial renewal and Wnt-dependent proliferation as a potential mechanism contributing to barrier dysfunction in IBD—complementary to established defects in adhesion. Failure to maintain or regenerate the epithelial barrier may be an important and under-recognised contributor to intestinal inflammation.

### Effector gene mapping informs drug prioritisation and safety for IBD

To explore how our effector gene nominations might inform therapeutic strategies, we first asked whether any of the colocalising genes were established or clinically relevant drug targets for IBD. We observed colocalisation between IBD risk and eQTLs for *ITGA4*, *JAK2* and *IL23R*, which are the direct molecular targets of vedolizumab and tofacitinib, respectively. We also identified colocalisations with *ITGAL*, *TNFSF15*, and *TNFRSF14* — genes that, while not directly targeted by existing therapies, participate in immune pathways modulated by current IBD drugs. Notably, the strongest regulatory effects for these genes were distributed across diverse tissues and cell types, including myeloid cells (*TNFSF15)*, plasma B cells (*ITGAL)*, and blood intermediate monocytes (*ITGA4)* (Supplementary Material S3). These findings are consistent with understanding that IBD treatments target a number of different immunological pathways, many of which are active across multiple cell types and tissues^61^.

We next asked whether our colocalised genes included targets of drugs approved or in clinical trials for other diseases, potentially suitable for repurposing. Using the ChEMBL database^62^, we identified several such candidates (Fig. 6). We observed colocalisation between UC risk and an eQTL for *PRKCB* (PPₕ₄ = 0.95, Supplementary Material S2), which encodes protein kinase C-β, a druggable kinase with inhibitors approved for oncology and already undergoing early-phase clinical trials for IBD. In our data, the IBD risk allele was associated with increased *PRKCB* expression in classical and intermediate monocytes, suggesting that heightened activity of this kinase may drive pathogenic inflammatory responses. Notably, PRKCB inhibitors are already undergoing early-phase clinical trials for IBD, and our findings provide human genetic support for their mechanism of action in relevant immune cell populations.

**Figure 6:**
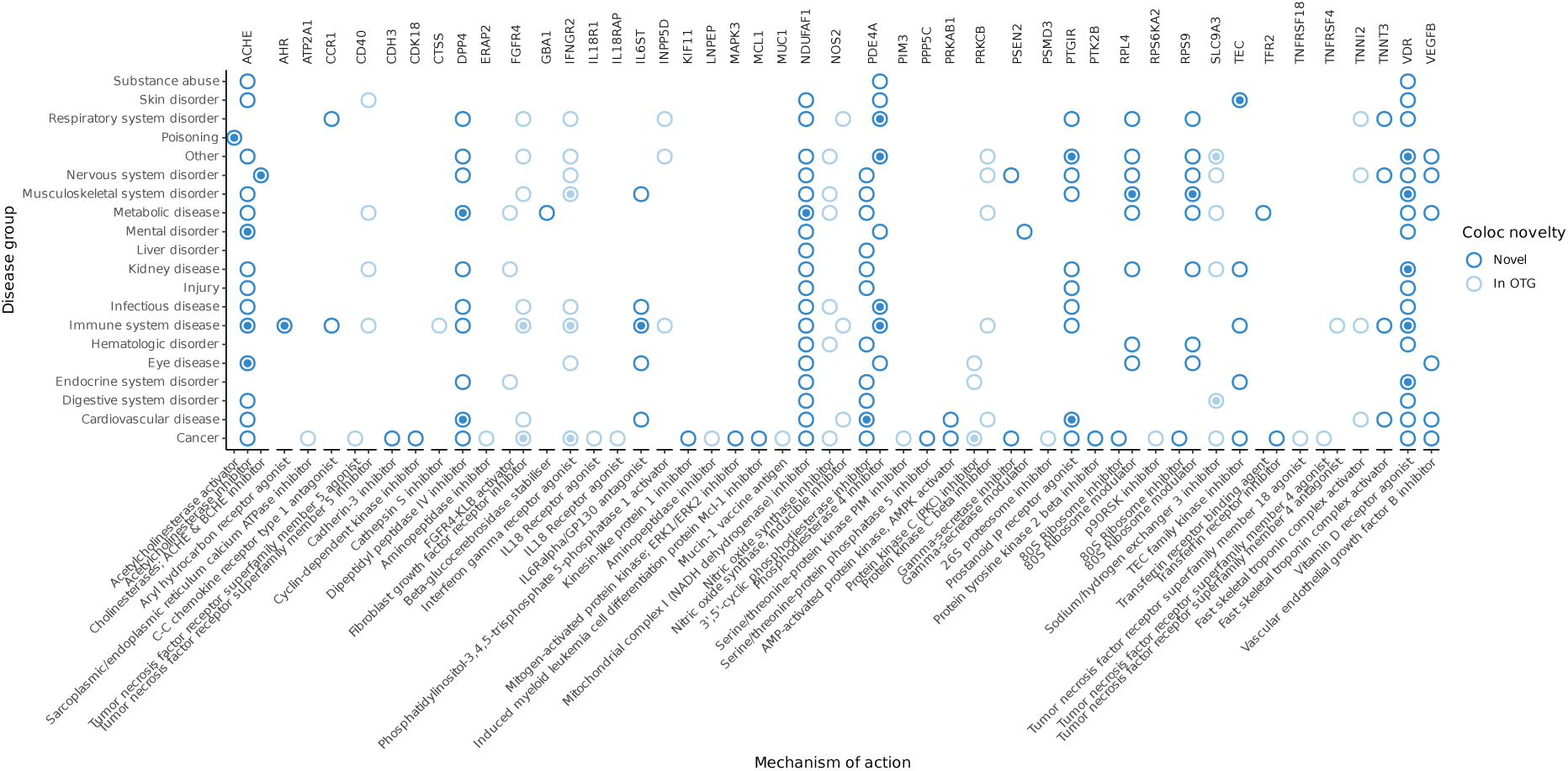
Identifying therapeutic repurposing opportunities by colocalisation. Comparison between the disease effector genes identified by colocalisation in IBDverse and the maximum stage of clinical trials therapeutic modifiers of each gene are described to be at in the CHEMBL database (see Methods). Hollow circle = maximum trial phase is 0.5-3, Circle with central dot = drug is approved for use/trial phase 4. Associations for genes that are previously colocalised in Open Targets Genetics (OTG) are light blue, genes that colocalise only in IBDverse are dark blue. Drugs are grouped by broad mechanism of action (x axis) and diseases are grouped into related families (y-axis)

We also explored whether our data could offer insights into potential on-target safety concerns. For example, we found colocalisation between a UC risk locus and an eQTL for *NDUFAF1* (PPₕ₄ = 0.88), which encodes a mitochondrial complex I assembly factor targeted by metformin which is approved for treatment of diabetes (Supplementary Fig. S11). The UC risk allele decreases *NDUFAF1* expression across many cell types, but the greatest explained variance in expression was in colonocytes, which may explain the common metformin side effects of diarrhoea, nausea, weight loss and abdominal discomfort^63,64^. Another notable example was *PSEN2*, where we observed colocalisation between UC risk and an eQTL (PPₕ₄ = 0.91) with discordant regulatory effects across cell-types (Supplementary Fig. S11). The risk allele was associated with elevated *PSEN2* expression in venous endothelial cells but decreased expression in classical and intermediate monocytes. *PSEN2* encodes presenilin 2, a catalytic component of the γ-secretase complex required for Notch receptor cleavage and activation. Mutations in *PSEN2* cause autosomal dominant early-onset Alzheimer’s disease by destabilising γ-secretase cleavage fidelity, leading to increased production of aggregation-prone Aβ42 peptides^65,66^. Although γ-secretase inhibitors were developed to reduce this pathogenic cleavage, their clinical use was curtailed by dose-limiting side effects such as weight loss and gastrointestinal bleeding^67,68^. Notably, murine *Psen2* knockout models also show susceptibility to colitis^69^. Our data suggest that *PSEN2* dysregulation in both immune and vascular compartments of the gut may underlie these adverse effects, and that inhibition of Notch signalling—while potentially anti-inflammatory in immune cells—may compromise epithelial homeostasis and barrier integrity. This context-specificity highlights the utility of eQTL colocalisation across diverse single cell atlases in anticipating tissue-specific effects of targeted therapies.

## Discussion

IBDverse represents the largest single-cell atlas of IBD-relevant tissues to date, encompassing nearly 2.2 million high-quality single-cell transcriptomes from 421 individuals, including 125 with Crohn’s disease. This size and diversity was critical for mapping eQTLs and nominating likely effector genes and cell-types shaping disease susceptibility. By including samples from Crohn’s disease patients with and without active inflammation, we captured regulatory effects in cell types that expand during disease but are rare or absent in healthy individuals—most notably myeloid subsets^70–73^, which harboured more IBD-colocalising eQTLs than any other major cell population. We also identify 764 ieGenes modulated by local inflammation, including two that colocalise with IBD GWAS at loci where an eGene did not. These context-dependent regulatory effects could only be uncovered by sampling inflamed tissue from individuals with active disease. At the same time, a large fraction of GWAS-colocalising eQTLs were identified in cell types abundant in healthy tissue, including epithelial, stromal and lymphoid populations. This underscores the power of generating cell-type-resolved eQTL maps in disease-relevant tissues even in the absence of disease. Given disease-affected tissues may not be accessible for some complex diseases, our findings reinforce the value of tissue atlases from non-diseased individuals for biological interpretation of GWAS loci — while also demonstrating the added insight gained by integrating samples from individuals with disease and relevant sub-phenotypic data.

A key insight from this study is the critical impact of cell-type resolution on eQTL discovery. While most eGenes were detected at the ‘All cells’ level (77.1%), an abundance of distinct regulatory variants (31.0%) were identified at the cell-type level only. Consistent with previous work^12^, eQTLs found at low resolutions were more proximal to transcription start sites and relatively enriched in promoters. However, cell-type level eQTLs were distal and more frequently located in enhancer regions—genomic features characteristic of GWAS loci. They were also less likely to regulate the nearest gene, suggesting more distal or context-specific modes of action. In line with these properties, we found that GWAS signals were substantially more likely to colocalise with eQTLs mapped at the cell-type level than with those detected at coarser resolutions. This suggests that cell-type level eQTLs represent regulatory effects that are more reflective of disease biology. More broadly, our findings imply that the genetic architecture of complex diseases is shaped by regulatory effects active in restricted cellular contexts—effects that are often missed in bulk analyses.

These properties are consistent with an evolutionary model in which regulatory variants that broadly affect the expression of disease-relevant genes are more likely to be deleterious and subject to negative selection. As a result, such variants would remain at very low frequencies in the population, limiting their detection in both GWAS and eQTL studies. In contrast, variants whose regulatory effects are restricted to specific cellular contexts may evade such selective pressures, allowing them to persist at higher frequencies and be more readily detected. This model provides a potential evolutionary explanation for the observed enrichment of GWAS loci among eQTLs mapped at the cell-type level.

Our findings have important implications for the design of future eQTL mapping studies aimed at identifying effector genes for GWAS loci. While we demonstrate the value of mapping at cell-type resolution, power to detect eQTLs was strongly correlated with the number of cells available per annotation. Despite a large overall sample size and nearly 2.2 million high-quality cells, 57% of our cell-type annotations had fewer than 100 cells per donor—for instance, none of our stromal populations had more than 25 cells per donor. This limits power to detect eQTLs, particularly for rare or condition-specific cell types—precisely the contexts in which disease-relevant regulatory effects may be missed by bulk eQTL mapping efforts. While we were able to nominate effector genes for over half of known IBD loci, it is likely that more comprehensive mapping in larger single-cell datasets, including samples from Crohn’s disease and ulcerative colitis patients with active inflammation, will enable effector gene discovery at a far greater proportion of loci. Such efforts would also support more accurate modelling of genetic effects on gene expression across cell types, allowing us to more precisely define the cellular contexts in which dysregulation contributes to disease.

Larger and more deeply powered single-cell eQTL studies will also enable more precise interpretation of regulatory mechanisms at individual loci, particularly where risk variants affect pleiotropic pathways across multiple cell types. Several of the loci we identified converge on the Notch signalling pathway, but do so through distinct mechanisms and in different cellular compartments. *MAML2* and *ZMIZ1* encode regulators of Notch-dependent gene transcription, and at both loci, reduced expression in dendritic cells colocalised with increased IBD risk. In contrast, findings at the *PSEN2* locus—encoding a catalytic subunit of the γ-secretase complex required for Notch activation—revealed opposing regulatory effects across tissues: the IBD risk allele was associated with decreased expression in monocytes but increased expression in endothelial cells. Together, these results support a model in which insufficient Notch signalling in immune cells contributes to IBD risk, while dysregulated or excessive Notch activity in non-immune compartments may promote gut toxicity. This duality may help explain the mixed outcomes of γ-secretase inhibition in clinical trials, where beneficial anti-inflammatory effects could be counterbalanced by epithelial or vascular side effects. These findings argue for therapeutic strategies that preserve Notch signalling in protective immune contexts while limiting its activation in others. More broadly, they underscore the value of cell-type-level resolution for dissecting the roles of pleiotropic pathways in complex disease.

A complementary mechanism suggested by our data involves disrupted epithelial renewal and stem cell maintenance. While barrier dysfunction is a well-established feature of IBD, prior models have largely focused on defects in epithelial adhesion. Our results point to an additional axis of risk involving impaired regeneration, with IBD risk alleles colocalising with eQTLs affecting genes such as *RASGRP1*, *MYC*, *LPIN3*, *RPS14*, and *FUBP1* that regulate Wnt signalling, epithelial proliferation, and mucosal repair. Many of these colocalised signals were assigned to colonocytes based on the proportion of gene expression variance explained by the eQTL, which was comparatively low in other epithelial subsets. Together, these findings motivate the hypothesis that disruption of epithelial renewal and stem cell maintenance in the absorptive epithelial cells of the rectum may be particularly relevant to IBD pathogenesis. However, the pathways involved are pleiotropic and tightly regulated, with well-documented roles in oncogenesis as well as repair. As such, while these results highlight the biological relevance of epithelial renewal mechanisms, they also underscore the challenge of safely modulating such pathways therapeutically. In this context, *FUBP1* provides a case in point: the same regulatory variant was associated with decreased expression in stromal cells—where it may impair regenerative programmes—and increased expression in immune cells, where it could enhance proliferation or inflammatory signalling. This complexity is further illustrated by an inflammation-dependent ieQTL for *RNF14*, a positive regulator of Wnt signalling, where reduced expression in inflamed enterocytes was associated with CD risk. These findings illustrate the complexity of cell-type-divergent regulation and reinforce the need for spatially and contextually resolved mechanistic studies to determine whether and how these pathways could be safely targeted in IBD.

This study demonstrates the power of single-cell eQTL mapping in disease-relevant tissues to resolve the cellular and regulatory architecture of complex diseases and traits. By systematically comparing genetic effects across cellular resolutions, we show that disease-associated variants are disproportionately enriched among eQTLs detected at the cell-type level, and often act through context-restricted regulatory mechanisms. These findings provide a framework for interpreting non-coding GWAS loci with greater precision, offer new insights into disease pathogenesis, and highlight the importance of sampling both healthy and inflamed tissues. As single-cell technologies continue to scale, the increased ability to map genetic effects at cell type resolution will enable more complete, mechanistically informed, and therapeutically relevant interpretations of complex disease association signals.

## Materials and methods

### Patient recruitment and single-cell sequencing

This study was approved by the National Health Service (NHS) Research Ethics Committee (Cambridge South, REC ID 17/EE/0338). Written informed consent was given by all participants.

296 individuals without inflammatory bowel disease (IBD) and 125 individuals with Crohn’s disease were recruited at Addenbrooke’s Hospital, Cambridge, UK. During routine endoscopy, gastrointestinal biopsies were collected from the rectum and terminal ileum, and blood samples were drawn after the procedure. Terminal ileal biopsies were obtained from 243 individuals without IBD and 119 individuals with Crohn’s disease. Rectal biopsies were collected from 275 individuals without IBD. Gastrointestinal biopsy processing was performed as previously described^70^, with an extended collagenase digestion step (15 minutes) for rectal samples due to their fibrotic nature. Whole blood was collected for single-cell RNA-sequencing from 95 individuals with Crohn’s disease. Peripheral blood mononuclear blood cells (PBMCs) were isolated from the whole blood using the EasySep Direct Human PBMC Isolation Kit (StemCell Technologies).

Cells were loaded for single-cell RNA-sequencing with a target recovery of 6000 cells per sample for IBD samples and 3000 cells per sample for non-IBD samples. Single-cell RNA sequencing was performed using 3’ 10X Genomics Chromium kits (v3.0 and v3.1) according to the manufacturer’s instructions. Libraries were sequenced to a target depth of 50,0000 reads per cell. CellRanger (v7.2.0) was used to align reads to the GRCh38 human genome reference with Ensembl v93 transcript definitions (GRCh38-3.0.0 reference file provided by 10X Genomics), and to generate cell-by-gene count matrices.

### Single-cell RNA sequencing quality control and cell clustering

Cell count matrices were initially subset to retain droplets likely to contain cells and corrected for ambient RNA using CellBender v2.1^74^, as previously described^70^. Scrublet v0.2.1^75^ was then used to identify and remove potential multiplets. All subsequent quality control, normalisation, and clustering steps were performed with Scanpy v1.9.6^76^, including normalising to library sizes of 10,000 total counts using the function sc.pp.normalise_per_cell and log-transformation using sc.pp.log1p to produce the ln[cp10k+1] count matrix.

A lineage-sensitive, relative QC strategy was applied to account for technical and biological variation across tissues and lineages (Supplementary Fig S1). First, the input dataset (4,099,804 cells) was aggregated and subjected to an initial round of quality control. Cells were removed if they had fewer than 250 genes expressed, fewer than 500 total counts, or more than 20% or 50% of reads originating from the mitochondrial genome, for blood-derived and non-blood-derived cells respectively. This left a total of 2,725,568 cells. Genes were then removed if they were not expressed in at least 5 cells. A total of 5,000 highly variable genes were then classified within each sample and merged using ‘sc.pp.highly_variable_genes’ and ‘flavour = seurat’. Ribosomal, mitochondrial, and immunoglobulin genes were then excluded from the highly variable set.

This data was then subject to an initial round of integration and clustering. Data was integrated using scVI v0.16.3^77^ function ‘scvi.model.SCVI’ on raw count data of highly variable genes. This was trained on 2 layers with 30 latent variables and gene likelihood modelled as a negative binomial distribution. A total of 50 nearest neighbours was calculated per cell using the function ‘sc.pp.neighbors’ to generate an adjacency matrix, which was used for calculation of both i) an initial UMAP embedding using function ‘sc.tl.umap’ with both a minimum distance and spread of 0.5 and ii) leiden clusters at a resolution of 0.03 using ‘sc.tl.leiden’. Initial clusters were then grouped into cell lineages, based on the expression of canonical marker genes (Supplementary Fig. S12), including; B cells, Epithelial cells, Mast cells, Mesenchymal cells, Myeloid cells, Platelets and T/ILC cells. Markers for these annotations are described in ‘*Annotation of cell clusters*’.

Within each cell lineage (B cells, Epithelial cells, Mesenchymal cells, Myeloid cells (not including Platelets or Mast cells) and T/ILC cells), we then further subset cells based on relative thresholds for quality control parameters. Cells with number of unique genes expressed, total counts and percentage of genes originating from the mitochondrial genome that lay outside the median plus/minus 2.5 median absolute deviations were removed. For the epithelial lineage, the median deviation was far greater than that for non-epithelial, and so a window of 2 median absolute deviations was utilised. Additionally, epithelial cells from samples with lower overall depth (indicated by median number of genes expressed per cell < 2000) were removed from the epithelial population only, and any infiltration of blood-derived cells were also removed. Highly variable genes were then re-selected from the full initial set within each lineage independently, and re-integrated using scVI as before, computing 20 nearest neighbours for non-epithelial cells and 50 nearest neighbours for epithelial cells.

Cell types were then identified by clustering within each lineage independently. As described previously^70^, the resolution at which final clusters were derived was determined in a data-driven manner. Cells from each lineage were clustered at resolutions of 0.1 to 1.5 (interval of 0.1) using the sc.tl.leiden function. To determine the reproducibility of the clusters detected at each resolution, two thirds of the data were then used to train a single layer dense neural network model using keras v2.4.3, and the per-cluster matthews correlation coefficient (MCC) of the fit was assessed in the remaining one third of data. After clusters that were either comprising >40% cells from a single sample or showed outlying quality control parameters were removed, we selected the resolution at which all remaining clusters had a MCC >0.75. Additionally, reproducible (MCC >0.75) clusters demarcated by the expression of genes indicative of poor quality cells, such as *MALAT1*^78^, and cross-lineage marker genes were also removed. The resolutions utilised for the final cell clusters were; Epithelial cells - 0.9, Mesenchymal cells - 1.0, Myeloid cells - 0.7, B cells - 0.8 and T/ILC cells - 1.1.

Finally, the additionally filtered and clustered per-lineage clusters were then recombined with the Mast and Platelet cells. This population, known as the ‘atlasing’ dataset, comprised 1,852,681 cells and was re-integrated as described above. With scope to auto-annotate cells with reliable transcriptional profiles that may have been removed by the strict, within-lineage quality control, a CellTypist v1.6.2^24^ model was trained on the ‘atlasing’ dataset. This was assessed by quantifying the proportion of cells that were correctly assigned to the same annotations in the initial, manual annotation. Overall, this was achieved at a rate of 97% (median 98%) (Supplementary Fig S13), with only 4 clusters achieving an accuracy <90% and only one <80% (Tissue Plasma B *CD38*^++++^ *IgG*^+^ *IgA*^+^ - 87.8%, Tissue Plasma B *CD38*^++^ - 79%, Blood atypical B cells - 88.7%, Tissue Memory B *HIVEP3^+^* - 80%). This model was then used to auto-annotate the full set of cells that passed the first round of quality control and QC, and filtered for those with confidence < 0.5, resulting in a final dataset that after intersection with genotyped individuals, results in 2,196,874 million cells.

### Annotation of cell clusters

As described above, cells from all tissues were initially clustered into cell lineages after the first round of quality control and integration. These were grouped and annotated on the basis of canonical marker genes (Supplementary Fig. S12); Epithelial cells were defined by high expression of *EPCAM*, *CDH1, KRT19*; Mesenchymal by expression of *COL1A1*, *COL1A2*, *COL6A2*, *VWF*; B cells by expression of *CD79A*, *MS4A1*, *MS4A1*, *CD79B*; Plasma B cells by expression of *MZB1, JCHAIN*; T and Natural killer (T/ILC) cells by expression of *CD3D, CD3E, CD3G, CCR7, IL7R, TRAC, NCAM1*; Myeloid cells by expression of *ITGAM, CD14, CSF1R, TYROBP*; Mast cells by expression of *TPSAB1*, *TPSB2*, *CPA3* and Platelets by expression of *RUNX1*, *GATA2*, *HDC* and *SLC24A3* (but low expressors of Mast cell genes). For within-lineage clustering, B cells and Plasma B cells were combined into a single lineage, with Mast cells and Platelets excluded.

Following clustering within lineage, cell types were first annotated by major population using a series of canonical markers, detailed in subsections below. In the case of T/ILC and mesenchymal populations, this matched their clustering lineage, however; Epithelial cells were further subdivided into Enterocyte, Colonocyte, Stem cells and Secretory cells; The B cell lineage was re-divided into B cells and plasma B cells and the myeloid cell lineage were combined with Mast and platelet cells. To interpret individual cell clusters, marker genes were calculated within major populations or across targeted subsets of clusters using scanpy’s sc.tl.rank_genes_groups function (method=’wilcoxon’). The significant results (FDR<0.05), along with a discriminative subset that were filtered using scanpy’s sc.tl.filter_rank_genes_groups (min_in_group_fraction=0.5, max_out_group_fraction=0.3, min_fold_change=1.5), were used to annotate and label clusters using expert knowledge. Enrichment of specific populations in a single tissue, or combination of tissues, was determined by the overall proportion of cells from each site (Supplementary Fig. S2).

#### Epithelial cell lineages

Genes used to demarcate epithelial lineages are presented in Supplementary Figure S14a. The stem cell major population was defined by expression of *LGR5*, *OLFM4*, *ASCL2*, *SMOC2* and *RGMB*. One of these populations was specific to the rectum, while two were specific to the TI, and hence are described as ‘Rectal’ and ‘Intestinal’ respectively. Within the intestinal subsets, one showed greater expression of *MKI67*, and the other with greater expression of *OLFM4* and *LGR5*, which are hence used to discriminate these populations.

Absorptive epithelial cells were also largely TI- or rectum-specific (Supplementary Fig. S2). The majority show clear separation in dimensionality reduced space (Fig. 1b), and were grouped into the Enterocyte (high expression of *RBP2, ANPEP, FABP2*) and Colonocyte (high expression of *CA2, SLC26A2, FABP1*) categories respectively. The one exception was a population composed of cells from both predominantly diseased TI and rectal origin, with high expression of *OLFM4*, hence referred to as “Epithelial progenitor *OLFM4^++^*”, grouped into the Colonocyte major population due to their embedding in dimensionality reduced space (Fig. 1b). Both Enterocyte and Colonocyte populations were characterised by the relative expression of marker genes, with discriminative ones being added to their label, including *OLFM4, MKI67* (indicative of proliferation), *BEST4*, *CEACAM7, KRT20, IFI27, ALDOB* and *RBFOX1*.

Secretory cells were broadly more shared across the TI and rectum, but with some site enriched clusters (Supplementary Fig. S2); Goblet cells were identified by expression of *MUC2*, *CLCA1* and *FCGBP*; Tuft cells by expression of *POU2F3*, *SH2D6, LRMP*, *TRPM5*; Paneth cells by expression of *DEFA5, DEFA6*; Enteroendocrine by expression of *NTS, PYY, GCG, ISL1* and Enterochromaffin by expression of *TPS1, CES1*.

#### Mesenchymal cell lineages

Markers used to demarcate mesenchymal cell subsets are displayed in Supplementary Figure S14b. Fibroblast clusters were identified by their expression of *ADAMDEC1, PDGFRA, BMP4, LUM, COL1A2, COL3A1* and *COL1A1*, subsets of which were then delineated by their expression of *pi16*, *C3, CPM, NPY, GRIN2A, ADAMDEC1* (lamina propria fibroblast), and tissue-specificity. Glial cells were identified by expression of *NRXN1*, Smooth muscle cells by expression of *ACTA2*, Pericytes by expression of *PDGFRB*, *RGS5* and *NDUFA4L2*, Endothelial cells by expression of *PECAM1*, *VWF*, *PLVAP*, *ACKR1* and *CD36*. Endothelial cells were further divided into inferred residency by expression of high *TLL1, ACKR1* and *PECAM1* (venous), modest *TLL1* and *PECAM1* (lymphatic) and high *CD36* greatest *PECAM1* and *PLVAP* (capillary). Myofibroblasts were identified by expression of *ACTA2* and *MYH11*, which were then delineated by relative expression of *GREM2*, *ADGRL3*.

#### B cell lineages

Markers calculated within Plasma B cells were not very discriminatory, however these clusters show a gradient of *CD38* expression, which is therefore outlined in the label (Supplementary Fig. S14c). Each of these include a majority of IgA expressing cells, with one cluster containing a minority of high IgG expressing cells. For the non-plasma B cells; Atypical B cells by expression of *FCRL2*, *ITGAX*, *TBX21*; Naive B cells by expression of *IGHD*, *FCER2*; Germinal centre/Plasmablast B cells by expression of *CD19, TCL1A* (for which we also identified a *MKI67* expressing, proliferative subset); Memory B cells by expression of *BANK1, FCRL2*. The several clusters of Memory B cells were also distinguished by the relative expression of marker genes; *CD27*, *HIVEP3, PTPRJ*, *PDE4D* and *TEX9*.

#### Myeloid cell lineages

Across the myeloid cell lineage, we identified several distinct clusters of monocytes, macrophage and dendritic cells (Supplementary Fig. S14d). Monocytes were defined by high expression of *S100A8*, *S100A9* and were broadly divisible by anatomical location. Blood monocytes were distinguished by expression of *CD14* and *FCGR3A* (CD16) and tissue monocytes by expression of *SOD2*, *CXCL9* and *CXCL10*.

Macrophages were predominantly identified in the TI/rectum and were identified by high expression of *ITGA4, CSF1R,* and *MAF*. Macrophage sub-populations could thereafter be distinguished by relative expression of *C1QA/B/C*, *CD163, MRC1, SELENOP,* and *HLA-DRA*.

Three separate DC subsets were identified: Type 1 conventional dendritic cells (cDC1s) were identifiable from expression of *CLEC9A* and *XCR1*, whilst cDC2s were identified by expression of *CLEC10A*, with plasmacytoid dendritic cells (pDCs) identified by *IL3RA*.

#### T and ILC lineages

Within the T and ILC lineage, T cells were defined by expression of *CD3D, CD3E, CD3G, TRAC*, *TRBC1, CD96*, refined by *CD4, CD8A*, *CD8B, CD3* and *GZMK* expression (Supplementary Fig. S14e). Naive subsets were identified by high expression of *SELL*; γẟ (GD) by expression of *TRGC2*, *TRDC* (for which we also defined gut- and blood-enriched subsets referred to as ‘Tcell_GD’ and ‘Tissue_Tcell_GD’, see Supplementary Fig. S2); Regulatory (T-regs) by expression of *FOXP3, CTLA4*; Effector memory subsets divided by expression of *TIGIT* and *THEMIS*); tissue resident memory with high expression of *TRGC2*. Two T cell clusters that did not express *CD8* or *CD4* were also identified, one of which strongly expressed *TRDC*, and the other that strongly expressed *GZMK*. Type 1 Natural Killer cells (NK1s) were identified by expression of *CX3CR1* and *FCGR3A*, and type 2 NK (NK2s) by *XCL1, XCL2* and *CD44*. Innate lymphoid cells (ILC) were identified by expression of *IL1R1, ALDOC, LST1* and *RORC*.

### Pseudobulking

Pseudobulk expression data was obtained by calculating the mean ln[cp10k+1] expression across cells of a given annotation, as per previous sc-eQTL mapping benchmarks^79^. This was performed at the All Cells, major population and cell type resolution, both within and across anatomical sites (Fig. 2a). Expression data was limited to samples with 5 or more cells in the given annotation and only taken forward for further analysis if there were 30 or more individuals contributing to the annotation. In addition, the genes present in each annotation matrix were limited to those with a total count greater than 0 in at least 20% of pseudobulked samples, resulting in a total of 24,761 unique genes being tested across all annotations. Pseudobulked expression matrices were then inverse normal transformed at the gene level prior to eQTL mapping.

### DNA collection and quality control

On the day of the endoscopy, blood (or gastrointestinal tissue, where blood was unavailable) was collected from individuals for DNA array genotyping. DNA was extracted from blood or tissue samples and genotyped with the Applied Biosystems UK Biobank Axiom Array in five batches of samples by YourGene Health. Genotypes were called jointly across all five batches of samples to mitigate potential batch effects. Genotype data were filtered to remove low missingness, duplicate samples, variants not in Hardy-Weinberg equilibrium, individuals with non-European ancestry (as predicted by KING^80^) and first-degree relatives. Variants were lifted over from human genome build 37 to human genome build 38 with LiftOver prior to imputation being performed using the Michigan Imputation Server with the TOPMed reference panel^81,82^. Imputed variants with imputation quality lower than 0.3 were removed.

### *cis*-eQTL mapping

Expression quantitative trait loci (eQTL) mapping was carried out using TensorQTL^83^ permutation and nominal modes with default parameters. *cis*-eQTLs were mapped for variants with minor allele frequency > 0.05 that were within a 2Mb window (1Mb in either direction) of the transcription start site of a given gene. The following model was used for mapping *cis*-eQTLs:

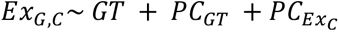

Where *Ex* is gene expression, *G* denotes a gene and *C* is an annotation. *GT* denotes the genotype alternative allele dosage of the individual (typed or imputed) and *PC* refers to principal components of genotype and annotation expression. To detect conditionally independent signals, a forwards regression framework was applied, as described previously^13^.

For interaction *cis*-eQTL mapping, a stricter MAF filter of 0.1 was applied to each variable level and an expanded model was used:

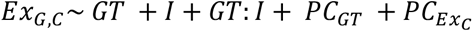

Where *I* is the interaction variable. We tested for interactions with sex, age (in bins of 5), disease status (CD or healthy), smoking status, and inflammation status (TI-SES-CD score, as defined previously^70^).

Five genetic principal components were included to control for population structure between individuals. The number of expression principal components to include was determined by repeating the analysis several times, increasing numbers of PCs from 2 to 100 in intervals of 2. The optimal model was selected based on the number of expression PCs that returned the greatest number of eGenes (FDR < 0.05), with ties broken by the model with fewest covariates. For the interaction analysis, any expression PCs which were correlated with the interaction variable were removed from the model (Pearson’s R > 0.25).

Lead variants for the same genetic association of a given gene may differ across conditions due to minor changes in significance, so to group lead variants across all tests and define eQTL signals, LD clumping was performed using the ‘--clump’ flag in plink v1.90b6.27^84^. Variants were clumped based on r^2^>0.5 with variants within 1Mb and with no additional significance filter.

### Colocalisation

Colocalisation was performed in a pairwise fashion between sc-eQTL summary statistics and GWAS summary statistics using coloc v5.2.2 and the coloc.abf function^85^. GWAS summary statistics from de Lange et al. (2017), were downloaded from GWAS catalogue (https://www.ebi.ac.uk/gwas/home), see ‘*Data and code availability*’. Analyses were conducted in 2Mb cis windows centred around the lead SNP of each eGene with a permuted FDR-corrected p-value < 0.05 and using GWAS loci which were significant at the genome-wide suggestive threshold (p-value < 5×10^−5^). Analysis was conducted using default prior probabilities of p1 = p2 = 10^−4^ and p12 = 10^−5^ where p1 and p2 are the prior probabilities of true genetic associations in each input dataset and p12 is the probability of true associations in both datasets. For colocalisation between GWAS and interaction eQTLs, a posterior probability of colocalisation (PP^h4^) greater than 0.75 was considered to be sufficient evidence for colocalisation of genetic signals. Colocalisations which fell within the HLA on chromosome 6 (build 38, positions 28,510,120 - 33,480,577) were discarded as likely false positives due to the complicated patterns of linkage disequilibrium in this region. Colocalisation events were assigned to the closest existing IBD locus, if the lead colocalised GWAS variant was within 500kb of a known IBD locus as defined by the interval comprising +/-500Kb containing: (i) the most likely causal variant from published fine mapping analysis^3,86^; or alternatively, if the locus has now been previously fine mapped (ii) from the gwas lead variant as defined in^3,28^.

To check if disease effector genes identified at high resolutions were more specifically expressed, genes were grouped into the lowest resolution at which a colocalisation was detected on their expression. The maximum specificity of gene expression at the cell-type level was then calculated using CELLEX^87^, and compared across resolutions using a wilcoxon test.

Detection of colocalisation events between two phenotypes is highly dependent on the significance of those effects. This is therefore underpinned by the statistical power in those tests, driven by the total number of observations. As this varies substantially for sc-eQTL tests (Fig 2b, Supplementary Fig. S3), inherent power to determine eQTL and colocalisation events also varies. To circumvent the influence of varied statistical power in the nomination of cell types within which colocalisation events are active, for each colocalisation, we quantified the proportion of variance in disease effector gene expression (r-squared) that was explained by the lead colocalising variant in each condition. This was calculated by linear regression of disease effector gene expression in the inverse normal transformed pseudobulk data on genotype.

### Enrichment of eQTLs in genomic annotations

To calculate the enrichment of lead variants from eQTLs in ENCODE and FANTOM genomic annotations, we followed previous approaches^12^, available on github https://github.com/hakha-most/gwas_eqtl. Briefly, eQTLs were first grouped by the minimum resolution at which they were detected, and their presence in functional elements was calculated. To calculate bootstrapped confidence intervals, we then sampled with replacement from independent LD blocks (calculated previously^88^) that contained eQTLs detected at each resolution one thousand times. Enrichment values in different annotations were then first normalised to that of random variants, before values in enhancers being normalised to those in promoters, in order to identify relative enrichment in these annotations specifically. Distal and proximal enhancer-like annotations from ENCODE were grouped together. P-values were calculated by two-tailed t-tests comparing the bootstrapped distributions, with 2.5-97.5% confidence intervals displayed in the enrichment plot.

### Therapeutic evidence analysis

We downloaded data on existing therapeutics and their clinical approval stage from ChEMBL and Open Targets^62,89^. Therapeutics were grouped into broad disease terms using the Ontology Lookup Service^90^ and filtered for those which directly targeted genes which we had colocalised with IBD (PP^h4^ > 0.75, within 0.5Mb of existing IBD locus) but had not yet been trialed as IBD treatments according to ChEMBL.

## Data and code availability

Raw single cell RNAseq data, processed expression data (both from the atlasing and eQTL mapping cohort), the CellTypist prediction model, genotype data, and summary statistics from all eQTL, ieQTL and colocalisation analysis will be made available upon publication. Code used to perform all quality control and clustering of scRNAseq data is available at https://github.com/andersonlab/atlassing. The pipeline used to perform QTL mapping is available at https://github.com/wtsi-hgi/QTLight, for colocalisation at https://github.com/andersonlab/snakemake_colocalisation, and for LD clumping at https://github.com/andersonlab/ld_clump_tqtl. Code used to generate all plots presented in this manuscript is available at https://github.com/andersonlab/IBDVerse-sc-eQTL-code. GWAS data is available from GWAS catalog (https://www.ebi.ac.uk/gwas/home) using accession numbers GCST004132 (CD), GCST004133 (UC) and GCST004131 (IBD).

## Acknowledgements

This research was supported by the NIHR Cambridge Biomedical Research Centre (BRC-1215-20014). The views expressed are those of the authors and not necessarily those of the NIHR or the Department of Health and Social Care. This research was funded in part by the Wellcome Trust [Grant numbers 206194 and 108413/A/15/D], The Crohn’s Colitis Foundation Genetics Initiative [Grant numbers 612986 and 997266] and Open Targets [OTAR2057]. We thank all individuals who kindly donated samples and their time to the study.

## 1 Supplementary figures

**Supplementary Figure 1:**
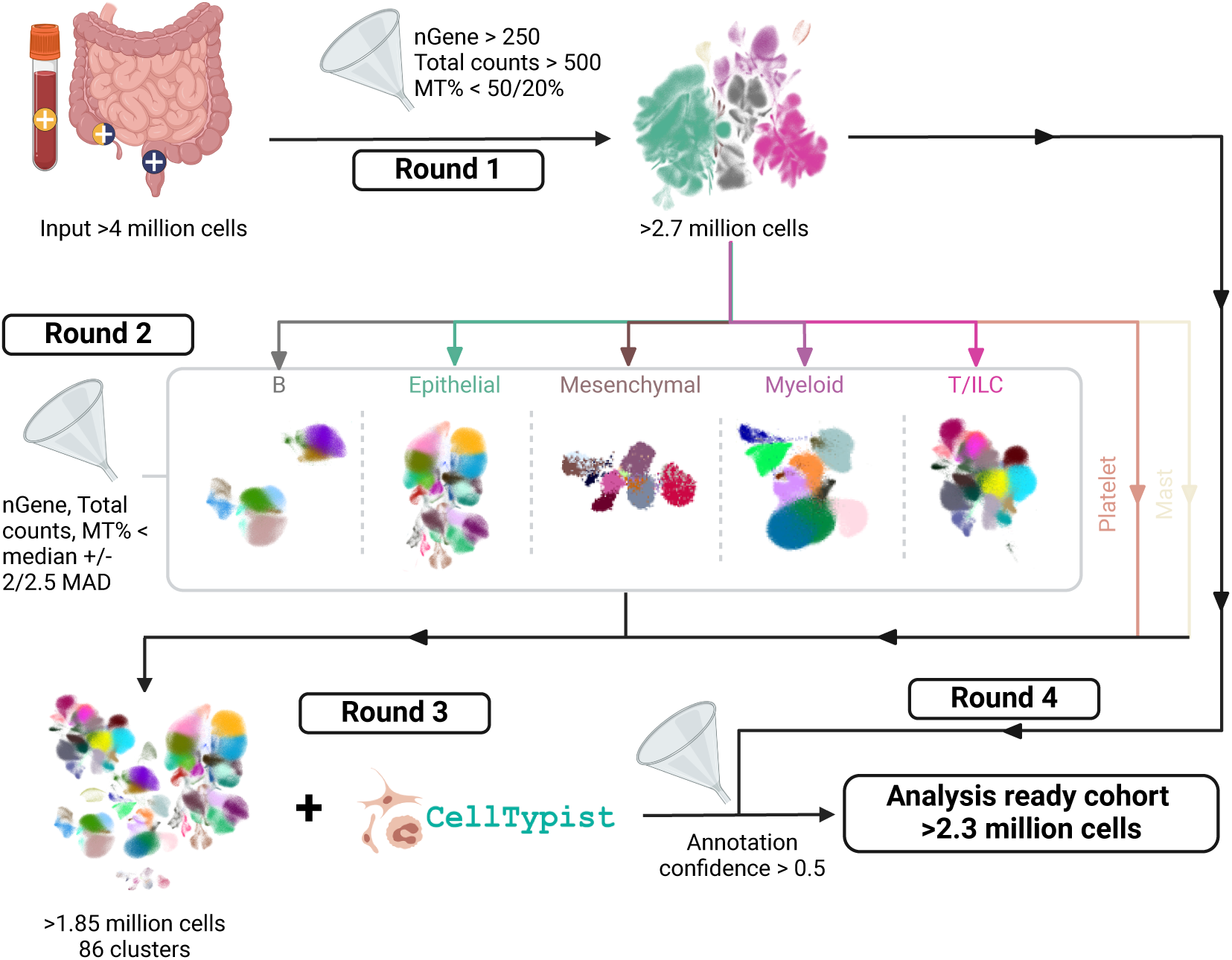
Overview of quality control, integration and clustering approach for scR-NAseq data. Analysis was performed across 4 rounds, a detailed summary of which is available in Methods. Cells were first filtered by basic quality control, integrated and clustered at a coarse resolution to identify broad lineages (see Supplementary Figure S12). These were then divided by lineage, and all except Platelet and Mast cells were subject to a second round of quality control using relative thresholds, before re-integration and clustering across a range of resolutions. The maximum resolution at which transcriptionally distinct populations of each lineage could be reproducibly obtained was then detected. Lineages were then recombined in a final round of integration, producing the total 86 cell-types. See Supplementary Figure S14 for outline of genes used to annotate individual cell-type. This was then used to derive a CellTypist model, which was used to annotate cells that passed quality control from the first round, producing the analysis-ready set. nGene = The number of genes expressed per cell, Total counts = total number of counts per cell, MT% = the percentage of counts mapping to genes in the mitochondrial genome, MAD = the number of median absolute deviations a cell’s quality control parameter is from the median for that given parameter.

**Supplementary Figure 2:**
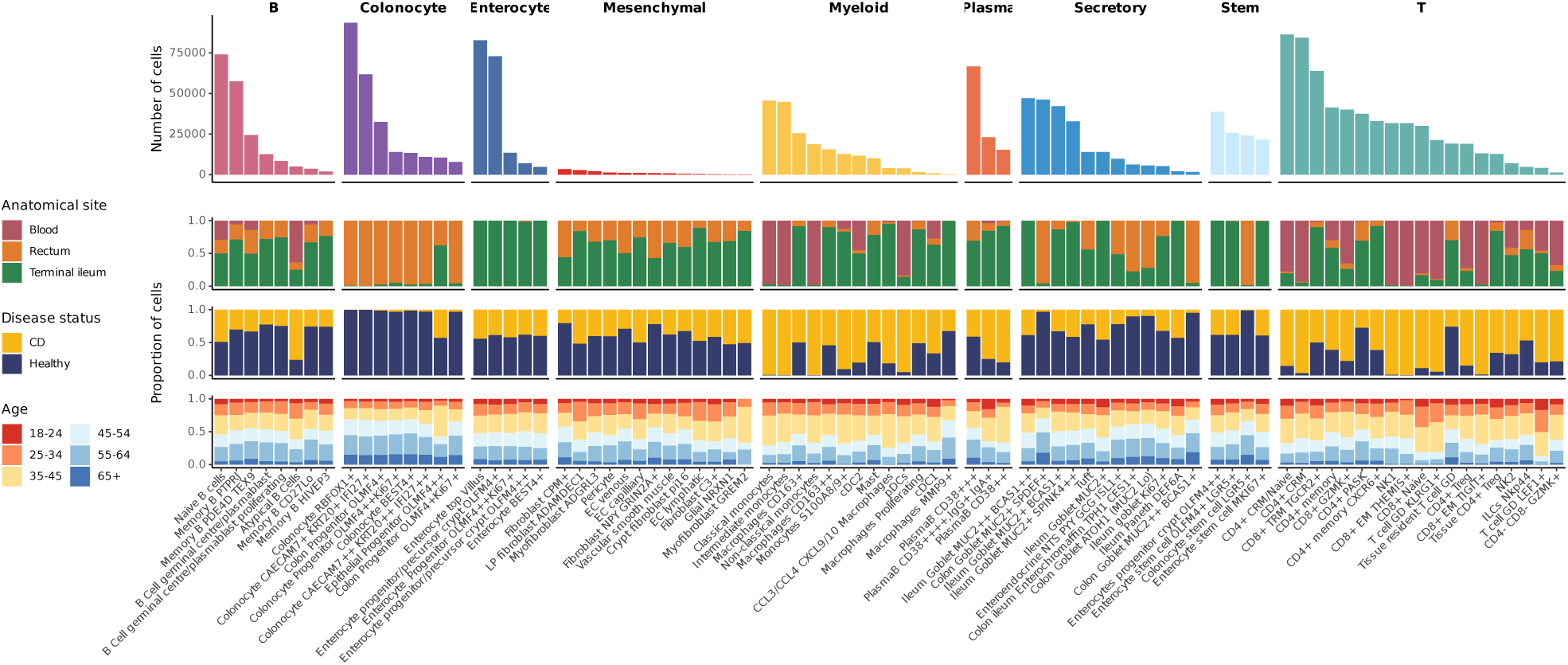
Cell-type level metadata. Total number (top) and contribution of different sites, disease status, and ages of sample to each cell-type in the 1,837,436 cells in the atlasing cohort.

**Supplementary Figure 3:**
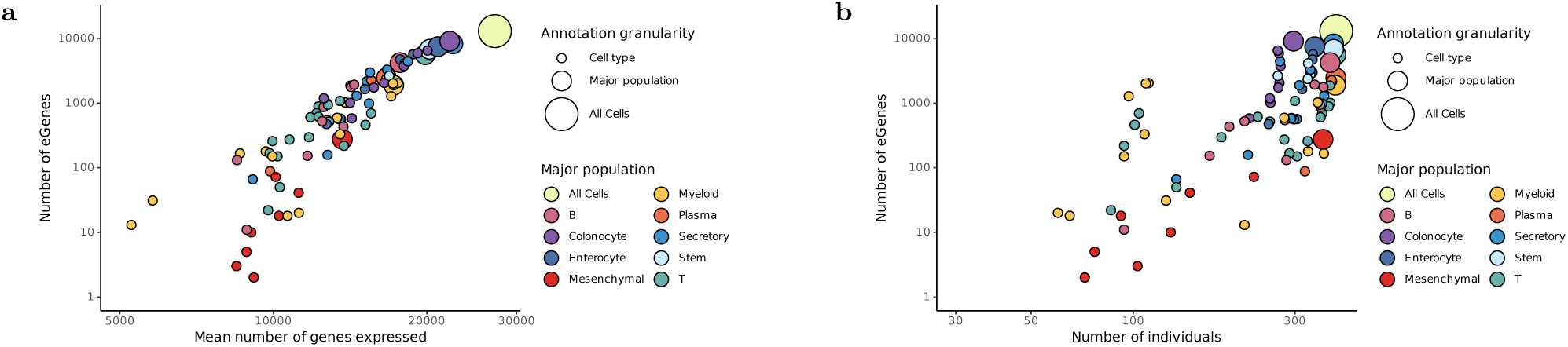
Factors influencing eGene detection. Association between between the number of eGenes detected in the cross-site analysis and a) the mean number of genes per pseudobulked sample or b) the number of individuals.

**Supplementary Figure 4:**
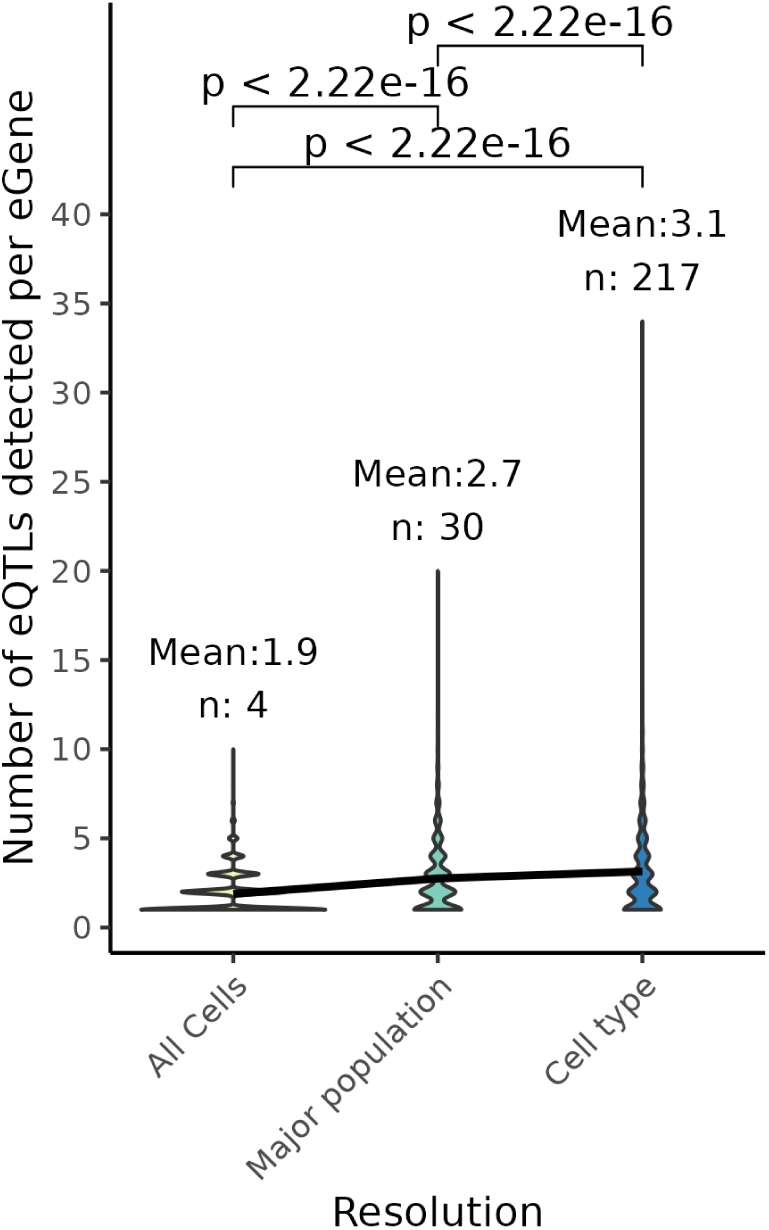
Number of eQTLs detected at each resolution. Comparison of the number of eQTLs (detectermined by linkage disequilibrium clumping, see Methods) that were found for each eGene at each resolution. Black line connects the means of each resolution, ‘n=’ indicates the total number of annotations per resolution, p-values calculated by wicoxon test.

**Supplementary Figure 5:**
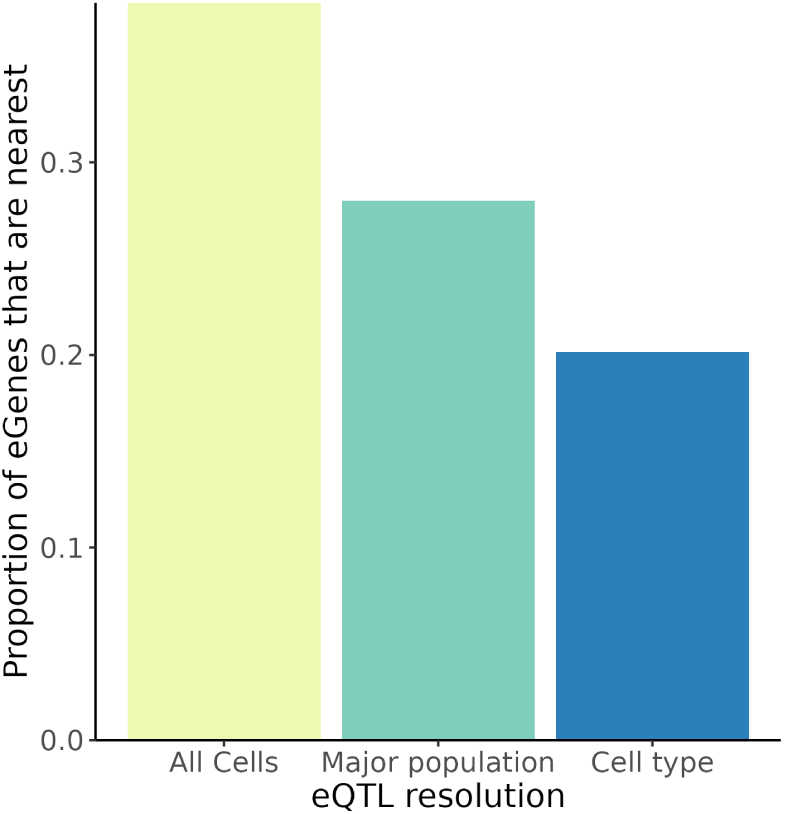
eQTLs detected at high resolutions are less frequently associated with the nearest gene. Proportion of eQTLs detected at each resolution for which the nearest gene is the eGene.

**Supplementary Figure 6:**
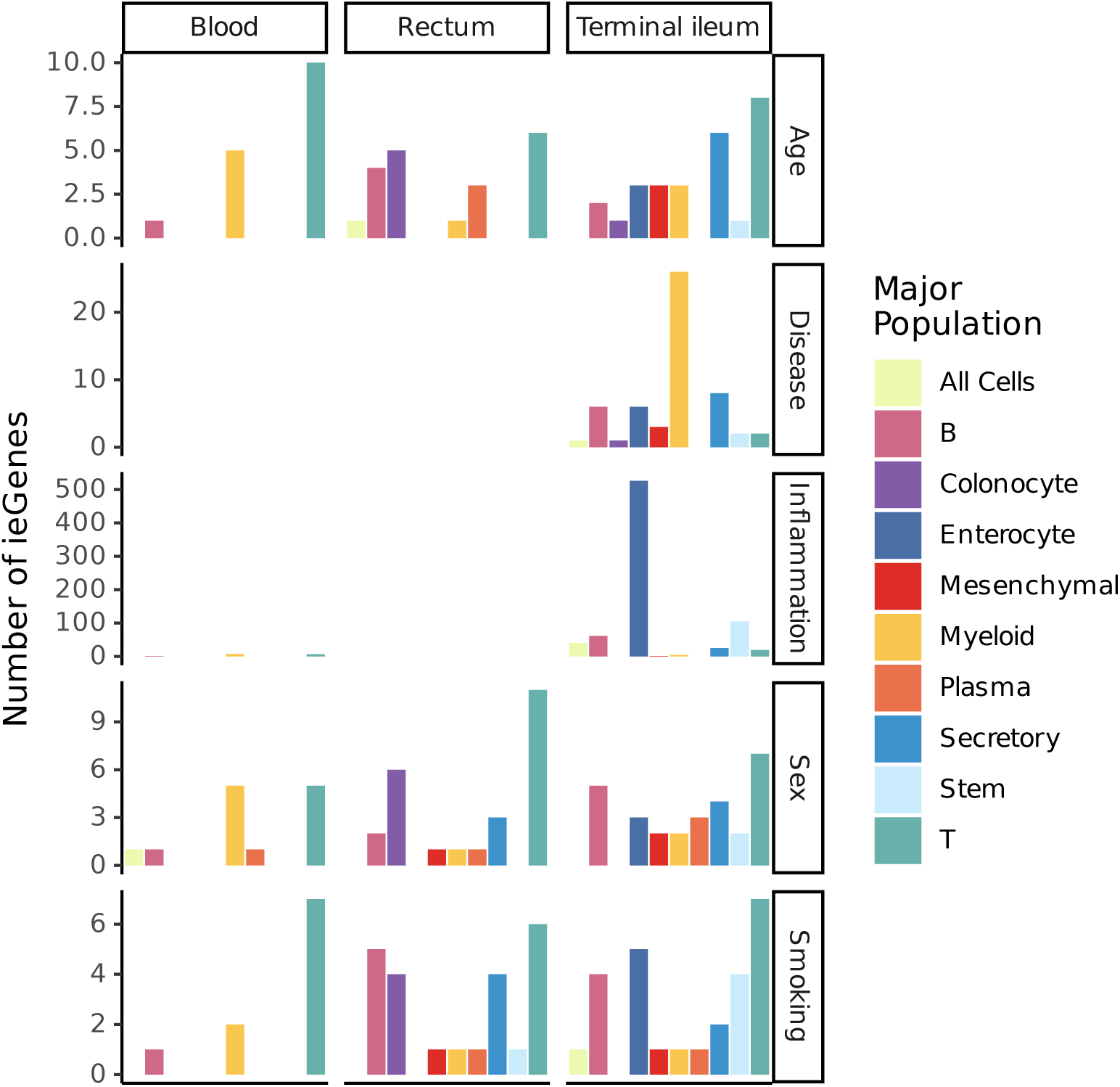
Number of ieGenes detected for each variable. Absolute number of eGenes detected by interaction (ieGenes) with sample level phenotypes, grouped by the interaction variable and the major population in which ieGenes were detected.

**Supplementary Figure 7:**
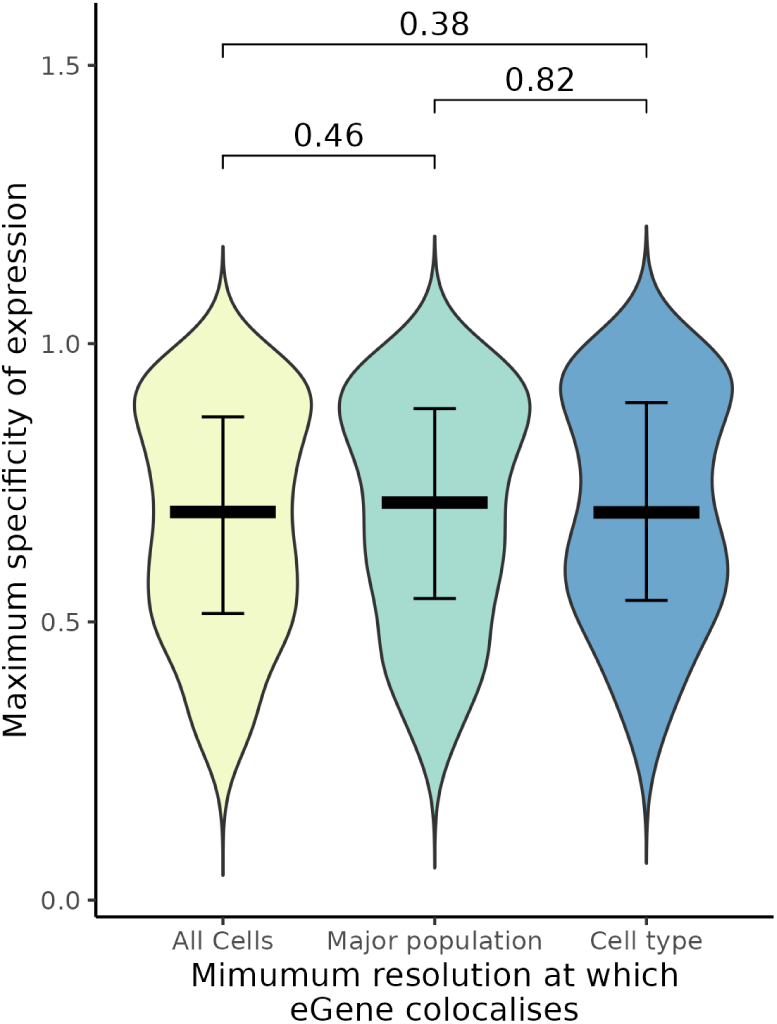
Disease effector genes identified at high resolutions are not more specifically expressed than those at lower resolutions. The maximum value of the per-cell-type expression specificity) of disease effector genes, as identified by CELLEX (Timshel *et al.,* 2022), grouped by the minimum resolution at which the colocalisation was detected.

**Supplementary Figure 8:**
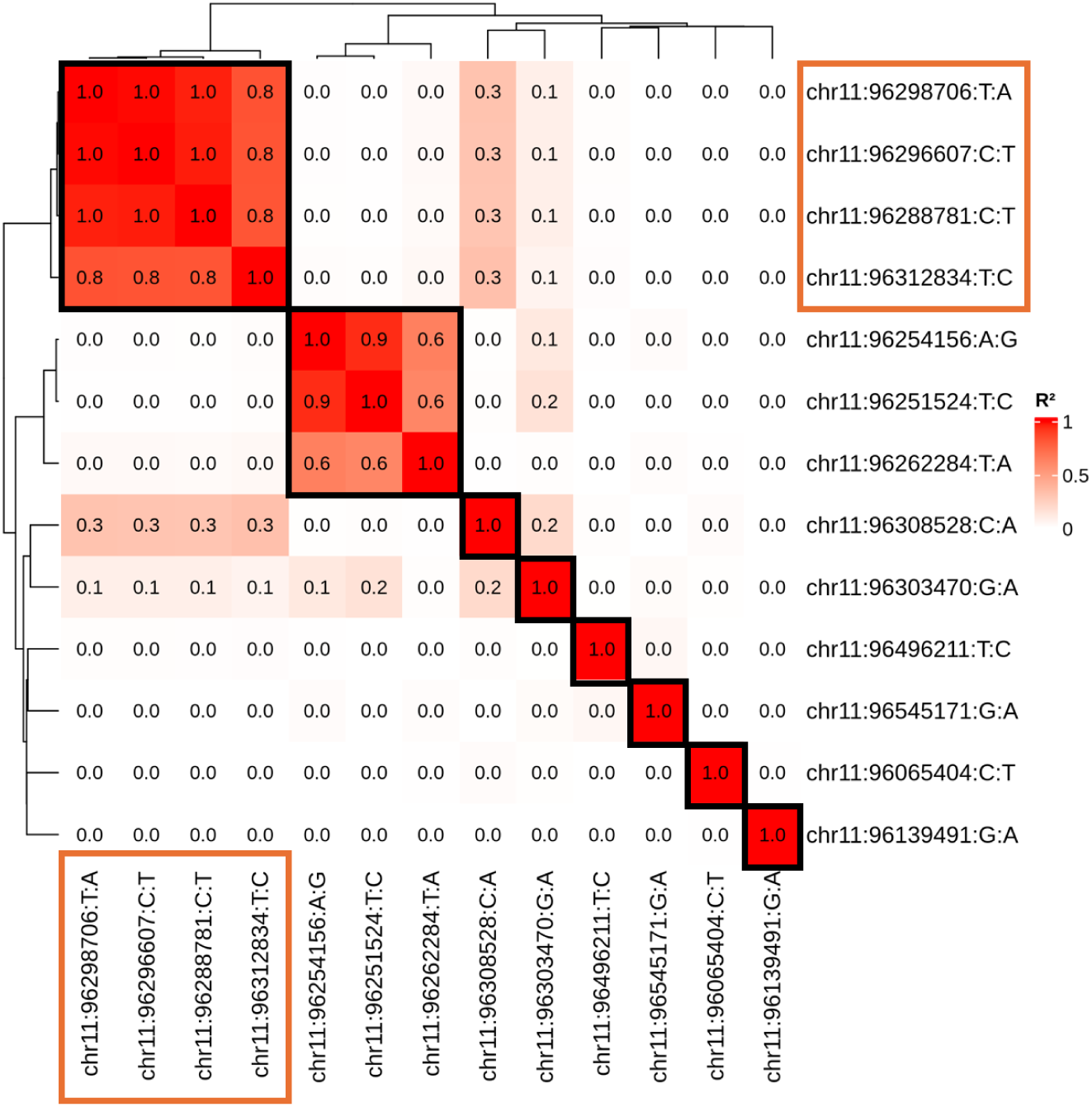
Independence of eQTLs for *MAML2* dysregulation. Heatmap depicting the pairwise linkage disequilibrium (r^2^) between lead variants from each condition with a significant eQTL. Clumped effects are indicated by black boxes around pairise tests. Lead variants that colocalise with IBD are outlined by an orange box.

**Supplementary Figure 9:**
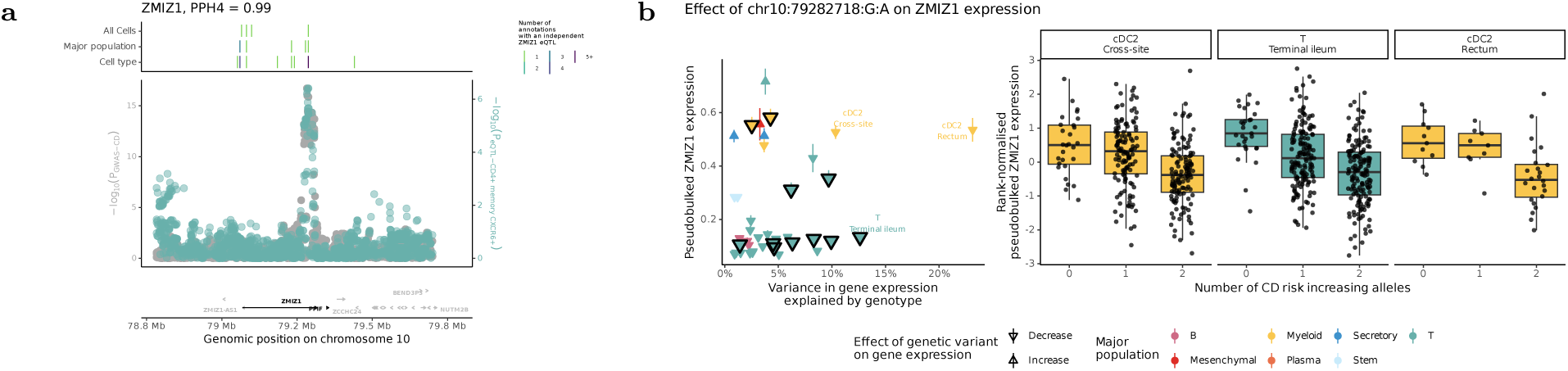
Identification and contextualisation of NOTCH signalling colocalisation target *ZMIZ1*. (a) Regional association pattern between genetic dysregulation of *ZMIZ1* and CD susceptibility. (b) Comparison of the variance of disease effector gene expression explained by the colocalising variant across conditions where a nominal (p < 0.05) eQTL effect was detected (left), and boxplots showing effect in the three annotations with the most variance explained (right).

**Supplementary Figure 10:**
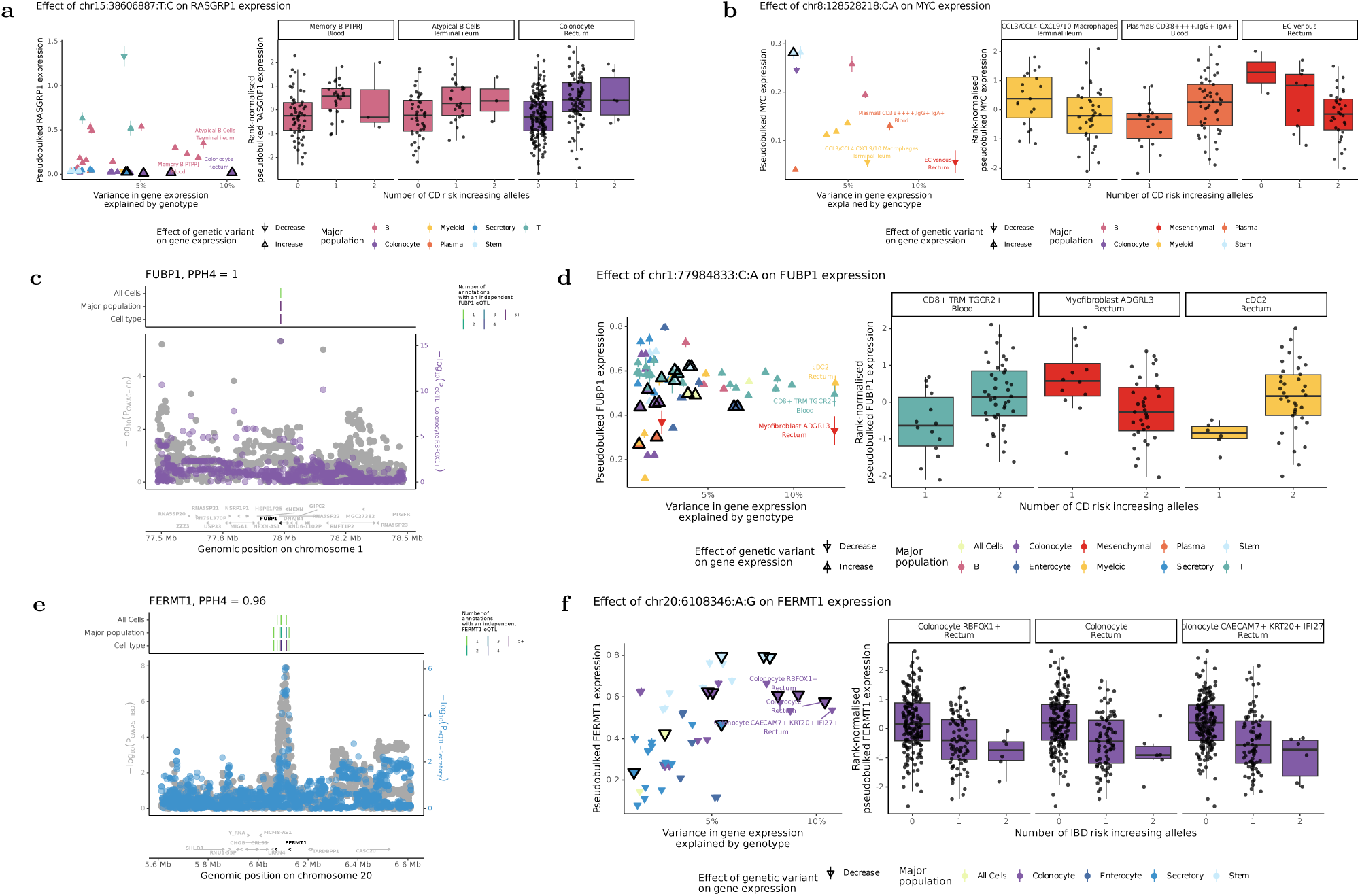
IBD colocalisations may impact epithelial barrier function and Wnt signalling. (a/b/d/f) Comparison of the variance of disease effector gene expression explained by the colocalising variant across conditions where a nominal (p < 0.05) eQTL effect was detected for each gene (left), and boxplots showing effect in the three annotations with the most variance explained (right). (c/e) Regional association pattern between genetic dysregulation of *FUBP1* and CD susceptibility or *FERMT1* and IBD susceptibility, respectively.

**Supplementary Figure 11:**
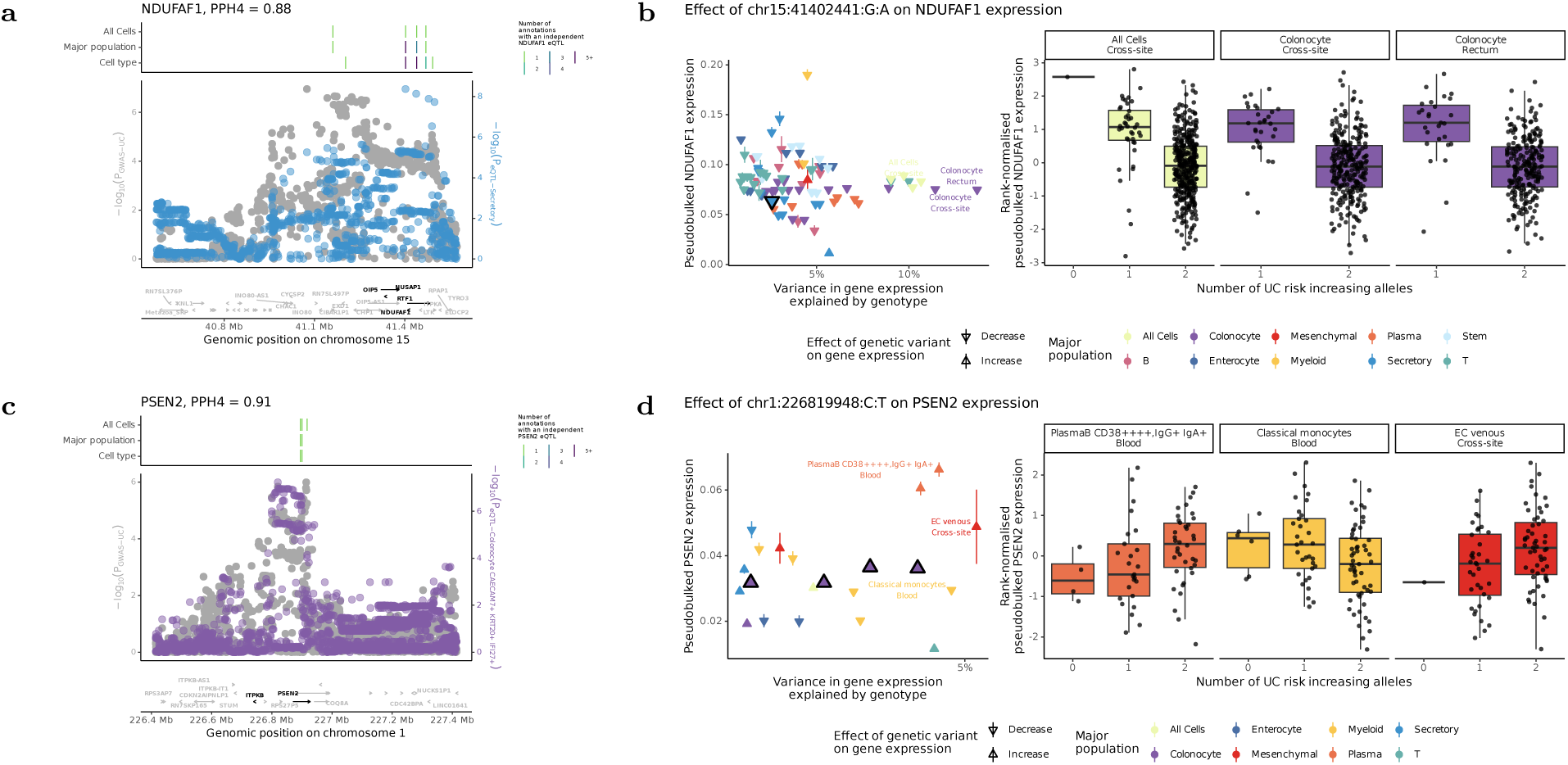
Colocalisation of UC with dysregulation of existing therapeutic targets *NDUFAF1* and *PSEN2*. Regional association pattern between genetic dysregulation of *NDUFAF1* (a) and *PSEN2* (c) and IBD types. Comparison of the variance of disease effector gene expression explained by the colocalising variant across conditions where a nominal (p < 0.05) eQTL effect was detected for each gene (b/d, left), and boxplots showing effect in the three annotations with the most variance explained (right).

**Supplementary Figure 12:**
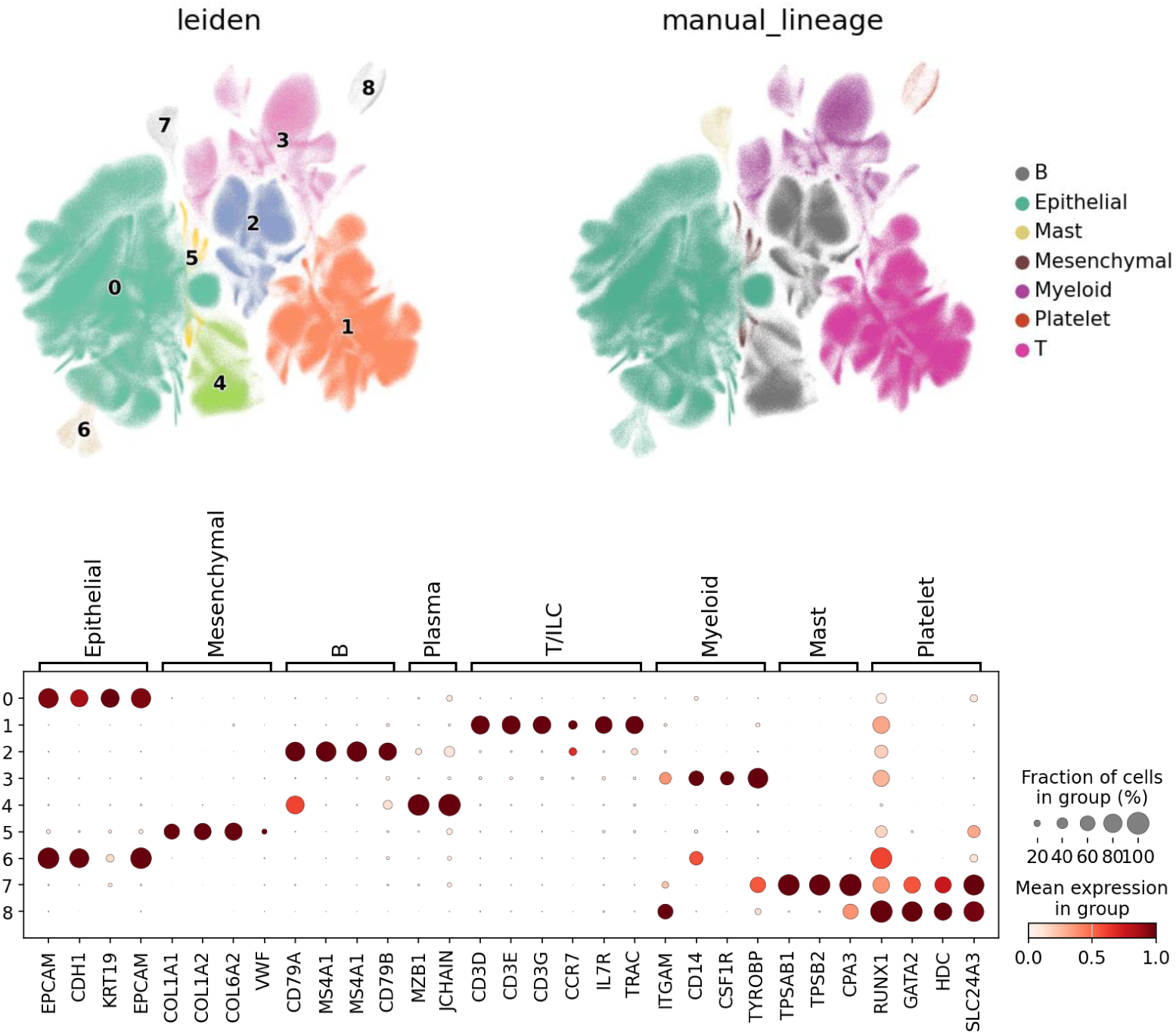
Identification of major lineages following round 1 of quality control and integration. Leiden clusters identified by clustering at resolution of 0.03 (upper left) following the first round of quality control and integration (see Supplementary Figure S1). Expression of canonical marker genes across leiden clusters, markers derived from literature (bottom). Annotation of the major lineages after grouping (upper right).

**Supplementary Figure 13:**
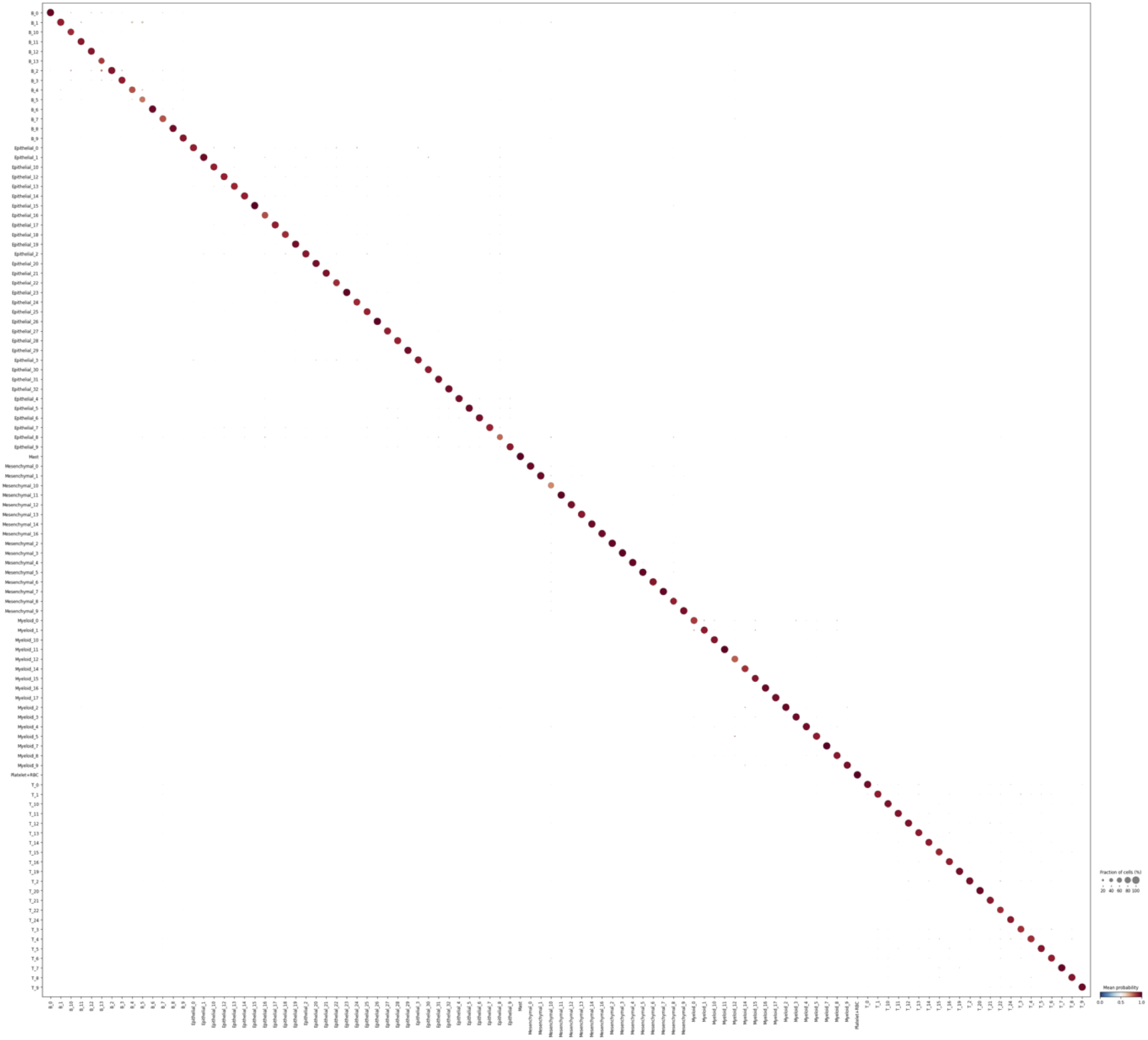
Accuracy of autoannotation model. Comparison of the original cluster labels (x-axis) with the CellTypist model predicted labels (y-axis) for cells in the original atlassing dataset.

**Supplementary Figure 14:**
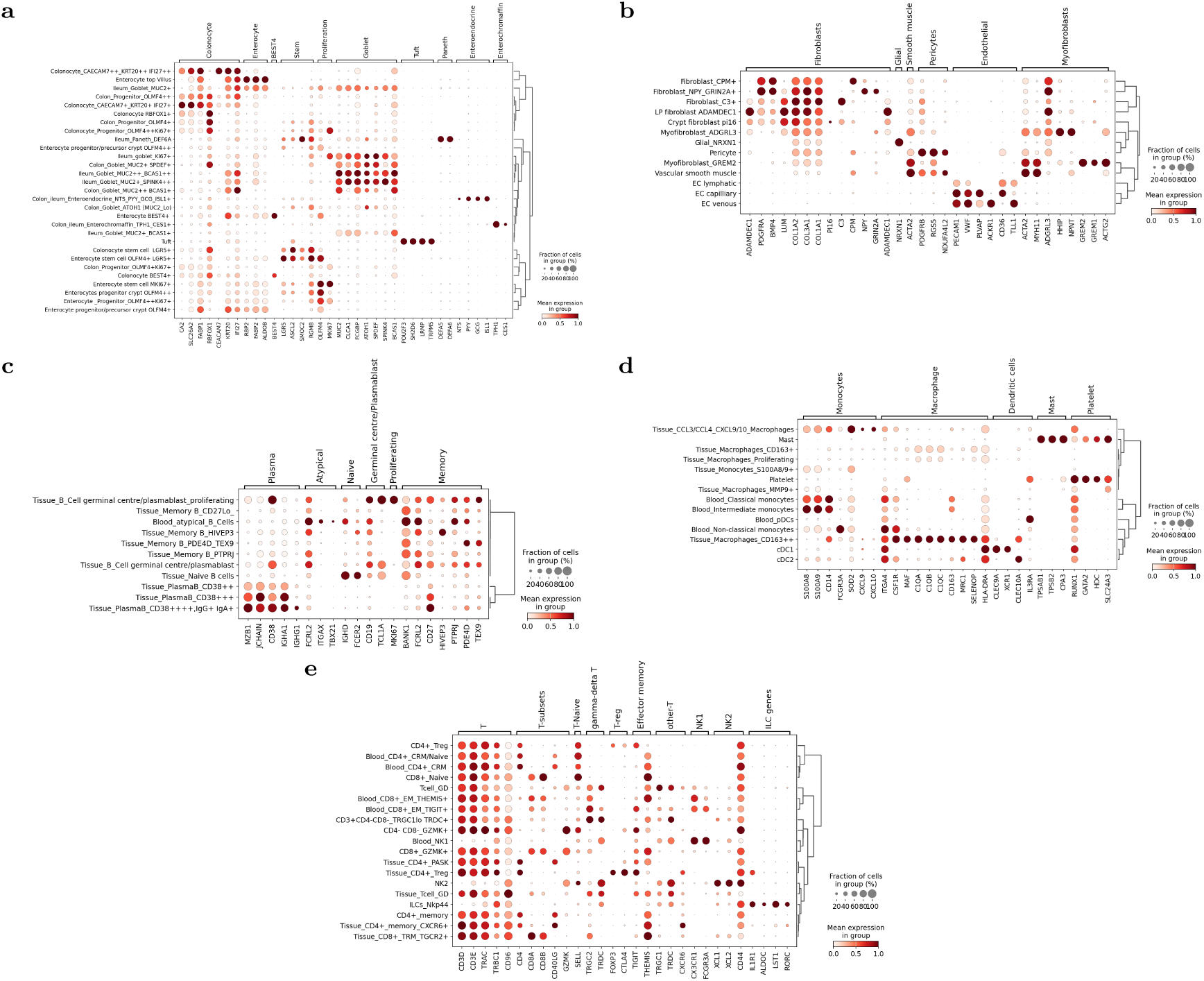
Marker gene expression for the cellular annotations. a-e) Dotplots for the expression of demarcating and discriminatory genes used to annotate clusters from each lineage, defined as in Supplementary Figure S12.

